# Diurnal modulation of subthalamic beta oscillatory power in Parkinson’s disease patients during deep brain stimulation

**DOI:** 10.1101/2022.02.09.22270606

**Authors:** Joram J. van Rheede, Lucia K. Feldmann, Johannes L. Busch, John E. Fleming, Varvara Mathiopoulou, Timothy Denison, Andrew Sharott, Andrea A. Kühn

## Abstract

**Background:** Beta band activity in the subthalamic local field potential (LFP) is correlated with Parkinson’s disease (PD) symptom severity, and is the therapeutic target and feedback signal for adaptive deep brain stimulation (aDBS). While clinically relevant beta fluctuations in PD patients are well characterised on shorter timescales and in the clinic, it is not known how beta activity evolves around the diurnal cycle, outside a clinical setting.

**Objective:** To characterise diurnal fluctuations in beta amplitude in PD patients receiving continuous, high frequency DBS.

**Methods:** We obtained chronic recordings (34±13 days) of subthalamic beta power in PD patients implanted with the Percept DBS device during high-frequency DBS, and analysed its diurnal properties. To investigate the influence of non-frequency-specific effects and artifacts, we compared beta to contralateral theta amplitude and recorded LFPs during various movements.

**Results:** Beta power had strong 24-hour periodicity, and time of day explained 41±9% of the variance in all long-term beta power recordings (p<0.001 in all patients). For all patients, beta activity was high during the day and reduced at night. Beta activity was not fully explained by theta activity and could show independent diurnal modulation. Movement artifacts affected the recorded LFPs, influenced band power estimates, and could have contributed to diurnal patterns in some patients.

**Conclusions:** Diurnal fluctuations in beta amplitude will need to be accommodated in aDBS to prevent suboptimal stimulation, particularly at night. Careful screening and/or mitigation of movement artifacts is needed to ensure that the signal is suitable for adaptive stimulation or neurophysiological investigation.

## Introduction

Subthalamic deep brain stimulation (DBS) is a highly effective therapy for Parkinson’s disease (PD) (Deuschl et al., 2006; Schuepbach et al., 2013). In addition to its clinical utility, DBS has provided unique insight into the pathophysiology of movement disorders, leading to the concept of abnormal network activity in brain disorders (Brown et al., 2001; Krauss et al., 2021; Kühn & Volkmann, 2017). The best characterized pathophysiological activity is an abnormal increase in the power and stability of beta oscillations in the neuronal activity of the subthalamic nucleus (STN) in the hypodopaminergic state (Brown et al., 2001; Hammond et al., 2007). Reduction of these pathologically enhanced beta oscillations correlates with the efficacy of treatment of akinetic/rigid symptoms using dopamine replacement or continuous, high-frequency (130Hz) DBS (Kühn et al., 2006, 2008). In the subthalamic local field potential (LFP), beta oscillations occur in “bursts” of high amplitude activity, the duration of which also relates to symptom severity (Tinkhauser, Pogosyan, Tan, et al., 2017).

These observations have inspired closed-loop, adaptive DBS (aDBS), where the amplitude of local beta band activity is used to adjust stimulation in response to pathophysiological activity (Arlotti et al., 2018; Bocci et al., 2021; Little et al., 2013; Swann, Hemptinne, Thompson, et al., 2018; Tinkhauser, Pogosyan, Little, et al., 2017). This approach has recently become more widely available through implementation on new DBS devices with the capacity to monitor beta power during continuous stimulation (Cummins et al., 2021; Feldmann, Neumann, Krause, et al., 2021; Neumann et al., 2017; Velisar et al., 2019). aDBS utilises changes in beta amplitude on the scale of seconds and minutes to control stimulation. However, in addition to the influence of stimulation amplitude and medication, beta power is likely to fluctuate in relation to processes on longer timescales. Most obviously, the occurrence of beta oscillations will be heavily influenced by sleep (Zahed et al., 2021). The balance of low to higher frequency activity in the STN LFP changes markedly from wakefulness to non-rapid eye movement (NREM) sleep, with higher delta and lower beta and gamma in PD patients (Thompson et al., 2018). Moreover, many neural processes are modulated by the output of the suprachiasmatic nucleus of the hypothalamus, which provides the brain’s principal circadian clock (Kim et al., 2019). This intrinsic circadian rhythm generator could influence beta oscillations both through its regulation of the sleep/wake cycle and the vast number of physiological processes that are influenced by changes in its output throughout the entire 24-hour cycle (Paul et al., 2020). Finally, levels of physical activity are also related to the time of day. This may lead to physiological modulation of beta in response to movements (Jenkinson & Brown, 2011; Kühn et al., 2004; Lofredi et al., 2019; Neuville et al., 2021; Quinn et al., 2015), but will also increase potential artifact sources. It is already known that the heartbeat can affect measures of beta power in certain DBS devices (Neumann et al., 2021; Sorkhabi et al., 2020), and movements themselves present another potential source of transient artifacts in LFP recordings (Hammer et al., 2021; Swann, Hemptinne, Miocinovic, et al., 2018). As the parameters for aDBS are currently set under controlled conditions during clinician working hours, both physiological and artifactual diurnal changes in beta measurement could impact aDBS effectiveness outside of the clinic.

Here we analysed long-term (18-59 days) recordings of STN LFP beta-band power from patients implanted with the Medtronic Percept DBS device, and show that beta amplitude can be significantly modulated according to time of day. We provide evidence that such fluctuations can be driven by physiological processes in some patients, but by movement-related artifacts in others. These findings highlight the importance of understanding and interpreting the sources of slow fluctuations in beta amplitude recorded from chronic sensing devices, for delivery of aDBS and for scientific investigation.

## Methods

### Participants

Written informed consent was obtained from all patients included in this study. The study was approved by the ethical review board of Charité Universitätsmedizin Berlin (EA2/256/60) and adhered to the tenets of the Declaration of Helsinki. 12 patients with advanced PD implanted with bilateral electrodes for STN DBS and a Percept IPG (Medtronic, Minneapolis, MN, USA) capable of chronic sensing participated in the study (surgery as previously described (Feldmann, Neumann, Faust, et al., 2021)). Patients 1-7 were implanted with model 3389 leads (Medtronic), and patients 8-12 were implanted with Medtronic SenSight electrodes (model B33005). Patients 1-6 (age 61.2±4.7 years; disease duration 10.8±3.7 years; 3 female) were included in the long-term bilateral beta monitoring data set, while patients 7-12 (age 65.3±7.5; disease duration 12.2±4.4 years; 1 female) were included in the simultaneous contralateral beta and theta power monitoring data set. Clinical severity and therapeutic effect were assessed using the MDS-Unified Parkinson’s Disease Rating Scale (UPDRS)-Part III. Prior to DBS implantation, motor impairment significantly improved with medication with average UPDRS-III change of 49±17%. Postoperatively, all patients benefited from chronic STN DBS with a mean improvement in UPDRS ON DBS/OFF medication of 43±16% (n=7, patients 8-12 have not yet passed the follow-up). Full clinical details are provided in Table 1.

**Table 1:** Clinical information and recording details: Information on the participants included in this study. Age at surgery, sex (F=female, M=male), disease duration in years. MDS-UPDRS Part III for OFF medication/ON medication state at the pre-DBS evaluation and during OFF Medication/DBS OFF/ON at 3 months follow-up for patients 1-6 and 12 months follow-up for patient 7. Patients 1-6 were included in long-term analysis, while in patients 7-12 beta/theta band activity was recorded simultaneously around the 12 months follow-up (patient 7) or in a subacute post-surgical phase (patients 8-12). Accordingly, LEDD are presented at the time of discharge from the 12 months follow-up for patient 7, after surgery for patients 8-12 and at the admission for three months follow-up for patients 1-6. Stimulation amplitude was either set to a single value or was allowed to vary within the given range during the recording period. Peak frequency for chronic sensing was set within the beta range on all STNs under investigation for patients 1-6, while for patients 7-12 one STN was recorded in the theta frequency range respectively. *: At 12 months-follow-up. **: Additional data collected to investigate beta power across sleeping and waking periods (not in main data set). ***STN excluded from analysis because of ECG contamination.

| ID | Age range (y) | Sex | Disease duration (y) | UPDRS pre-DBS OFF/ON MED | UPDRS OFF/ON DBS, OFF MED at 3-months follow-up | Time of recording (days post-surgery [duration]) | Stimulation amplitude (mA) | Peak frequency (Hz) | LEDD at time of recording |
| --- | --- | --- | --- | --- | --- | --- | --- | --- | --- |
| 1 | 61-65 | F | 10 | 50/9 | 22/15 | 43-74 (31) | R:2.2-2.8<br>L:0.1-0.2 | R: 25.39<br>L: 19.53 | 575 mg |
| 2 | 66-70 | M | 11 | 63/30 | 48/34 | 123-155 (32) | R:0.5<br>L:0.5 | R: 27.34***<br>L: 16.60 | 1175 mg |
| 3 | 56-60 | F | 15 | 37/23 | 29/20 | 32-64 (32) | R:1.4-1.6<br>L:1.4-1.8 | R:25.39<br>L:13.67 | 100mg |
| 4 | 61-65 | F | 6 | 50/33 | 40/19 | 73-91 (18) | R: 1<br>L: 1 | R: 14.65<br>L: 18.55*** | 300 mg |
| 5 | 66-70 | M | 15 | 66/27 | 39/14 | 37-96 (59)<br>+21** | R:2-2.8 | R: 15.63 | 300 mg |
| 6 | 56-60 | M | 8 | 23/4 | 28/11 | 116-148 (32) | R:1.7-2<br>L:1.7-2 | R:22.46<br>L: 20.51 | 175 mg |
| 7 | 56-60 | M | 11 | 72/39 | 47/33* | 393-396 (3) | R: 1.2<br>L: 2 | R: 20.51<br>L: 6.84 | 711 mg |
| 8 | 66-70 | F | 11 | 49/30 | N/A | 10-20 (10) | R:0-1<br>L:0-1 | R: 23.44<br>L: 7.81 | 580mg |
| 9 | 71-75 | M | 15 | 67/43 | N/A | 8-19 (11) | R: 0.5-1.5<br>L: 0.5-1.5 | R: 15.63<br>L: 7.81 | 575mg |
| 10 | 66-70 | M | 18 | 40/25 | N/A | 8-14 (6) | R:0.5<br>L:0.5 | R: 7.81<br>L: 13.67 | 800mg |
| 11 | 66-70 | M | 5 | 24/13 | N/A | 3-5 (2) | R:0<br>L:0 | R: 7.81<br>L: 15.63 | 832mg |
| 12 | 56-60 | M | 13 | 68/39 | N/A | 10-14 (4) | R: 1<br>L: 1 | R: 7.81<br>L: 13.67 | 900mg |

### Sensing

Frequencies for chronic recording were visually selected in power spectra generated by the BrainSense signal test, which automatically computes a fast Fourier transform of 20-second LFP recordings from the possible bipolar recording configurations per hemisphere. The recording contacts with the most prominent beta peak were selected. A window of 5Hz around this peak was used for long-term sensing of peak power (µVp) as an average every 10 minutes, with associated time stamps in co-ordinated universal time (UTC). As data were collected in Germany in two different time zones (Central European Time and Central European Summer Time), time stamps were corrected to reflect local time. In patients 1-6, beta power was collected for an average of 34±13.4 days at a stable optimized medication and stimulation regime until the 3-months follow up. Patients 7-12 were included for the data set exploring frequency specificity of the beta band fluctuations in subacute recordings after DBS surgery (duration 6.0±3.7 days). Peak beta frequency was selected as described above in one STN, while in the contralateral STN peak power was logged in a 5Hz window around a theta frequency (7.65±0.40Hz) for 6.0±3.7 days, irrespective of whether an oscillatory peak was present. Medication and stimulation amplitude were adjusted as per standard of care. Stimulation amplitudes and sensing frequencies for all patients are provided in Table 1.

### Assessing the impact of movements and medication on the LFP signal

Additional recordings during different types of movement were performed in patients 3, 4, 5 and 6 while they were on their usual medication. LFPs were recorded using the Percept IPG, sampled at 250 Hz during DBS, streamed to the Medtronic clinician programmer, and exported to the json-file format. After a rest recording of 3 min in a sitting position, patients were asked to perform head movements (nodding (24±10.98s), shaking (24.63±8.81s), tilting to the sides (27.63±10.31s)), arm movements (swinging back and forth (29.86±2.84s), lifting the arms to the sides (30.36±3.5s), closing the hands in front of the chest (36.25±18.37s)), and axial movements (chest rotation (29.13±2.84s), bending backwards and forwards (31±10.12s)). Patients also changed position (sitting (15.92±6.00s), standing up (2.25±0.40 sec), standing (15.79±6.57s), and sitting down (2.71±0.75s), at least 3 times) and walked back and forth (walking (26.13±4.13s) and turning (2.63±0.25s)). Video recordings of the patients were registered to the LFP signal by videotaping the start of each LFP recording on the programmer tablet. Subsequently, time stamps for each movement were aligned to the LFP signal. Any ECG artifacts were detected and removed from the LFP streams using the Perceive toolbox (Neumann et al., 2021). Signal power estimates were calculated after application of a low-pass filter at 98Hz and a high-pass filter at 4Hz for each movement epoch type, with the beta band set as a 5Hz window around the STN-specific beta peak used for long-term sensing, while theta band estimates were calculated as band power between 4-8Hz. For the comparison of in-clinic daytime beta power with the out-of-clinic long-term measurements, additional 60-second at-rest LFP recordings were obtained from patients 1 and 3 after at least 12 hours withdrawal of dopaminergic medication and after controlled administration of fast-acting levodopa (100-200mg L-dopa 30+ minutes prior to recording), on and off DBS.

### Data analysis

Analysis was carried out using custom-written scripts in MATLAB (Mathworks, Natick, MA, USA) and with the ‘Circa Diem’ analysis toolbox developed by author JJvR (v0.1, available on https://github.com/joramvanrheede/circa_diem, <u>DOI: 10.5281/zenodo.5961105</u>). ECG artifact influences were examined using the open source Perceive Toolbox ((https://github.com/neuromodulation/perceive/ (Neumann et al., 2021)). Unless otherwise stated, means are reported as mean±standard deviation and compared using (paired) two-tailed t-tests.

#### Detection of ECG artifact contamination and cleaning of LFP streams

The LFP streams from the BrainSense signal test for beta peak detection were screened for contamination by cardiac signals using the Perceive toolbox (Neumann et al., 2021). If an ECG artifact was identified by the classifier or observed on visual inspection, the STN long-term data were excluded, resulting in the removal of 2 STN time series (patient 2 right STN; patient 4 left STN). The ECG artifact analysis results for all 6 long-term bilateral sensing patients are included in Appendix 1, Supplementary Figure S1-6, A-C.

#### Outlier removal, data normalisation and de-trending

The LFP band power measure contained occasional extreme values. As the Percept device was only able to log a band power summary measure and not the raw data, inspection or recovery of the underlying LFP was not possible. We therefore removed outliers by replacing any values with a z-score greater than 6 with a value obtained from linear interpolation between their nearest neighbour values. This process was repeated until the data set contained no values with a z-score greater than 6. The mean number of outliers per data set was 18±23, and the maximum number of outliers for a timeseries was 71 (∼0.82% of data points over 59 days). For visual comparison between patients and hemispheres, long-term sensing data are sometimes presented as z-scores. For visualisation of diurnal patterns in heatmaps and rose plots, and to investigate diurnal correlations of between beta and theta power while removing any influence of longer-term drift from the correlation measure, time series are de-trended with the Circa Diem toolbox where indicated. De-trending was accomplished by dividing all LFP power measurements for a given day by that day’s median value (such that in the detrended data, the median for each day is 1).

#### Time-of-day fits, variance explained by time-of-day, and temporal shuffling test

To estimate the effect of time of day on the variance of LFP power time series, we created a fit based on the average values for each time of day. The day was divided into 30-minute segments, and the mean for this time bin was used as the fit estimate. The fit estimate for any value between the centres of time bins was then the result of a linear interpolation. The influence of time-of-day on the signal could now be ‘removed’ by subtracting the fit value for each time of day from the observed values. The variance explained by time of day was then calculated by taking the total variance of the time series, subtracting the variance remaining after influence of time-of day was removed, and then dividing by the total variance. To estimate how likely it was that a given value might be observed by chance, we devised a temporal shuffling test: we generated 1000 shuffled data sets by applying random shifts between 0 and 24h to the data for each day in a data set such that days were offset relative to each other but the time series for each day maintained its temporal structure (values that exceeded 24h were moved to the beginning of the day). This resulted in a null distribution of values for variance explained by time of day, to which the values obtained from the real data set could be compared. Time-of-day fits, the calculation of variance explained by time-of-day, and the temporal shuffling test are all implemented in the Circa Diem toolbox.

#### Estimates of sleeping and waking times and aligning data to estimated rise time

To examine beta power changes across periods of sleeping and waking, we acquired 21 days of additional bilateral STN data from patient 5, who was asked to log their bedtime and rise time each day. We approximated wake up time in other patients by aligning their data to their first scheduled medication intake. To estimate the effect of time of day during sleeping or waking hours only, we separated estimated sleeping and waking epochs for all patients during the long-term data acquisition by selecting a time window usually dominated by sleep (00:00-06:00) and a time window usually dominated by waking (08:00-20:00). We used the same time windows to obtain distributions of long-term STN LFP power values during estimated day or night-time, for comparison with the daytime in-clinic measurements on and off medication and DBS.

## Results

### Measured STN beta power fluctuates in a 24-hour cycle and is low during the night

In 6 patients implanted with bilateral STN DBS electrodes, we acquired measurements of beta oscillatory power of the LFP every 10 minutes. An LFP stream, power spectrum, and long-term beta power measurements are shown for a single STN in Figure 1A-C. Alternating periods of high and low beta power are clearly visible in the long-term measurements, and beta power was consistently higher during the day and lower during the night (Figure 1D-F). Indeed, all STN time series showed a clear 24h periodicity (Figure 1G), suggesting that the diurnal cycle had a strong effect on beta power. Time-of-day fits (such as in Figure 1E) explained a significant proportion of the variance in beta power in all time series (Figure 1H; mean 0.41±0.092; p<0.001, temporal shuffling test). Moreover, all STN LFPs showed a pattern of high beta power during the day and low beta power during the night (Figure 1I), though there was some individual variability in diurnal beta profiles (Appendix 1, Supplementary Figure S1-6, D-F).

**Figure 1:**
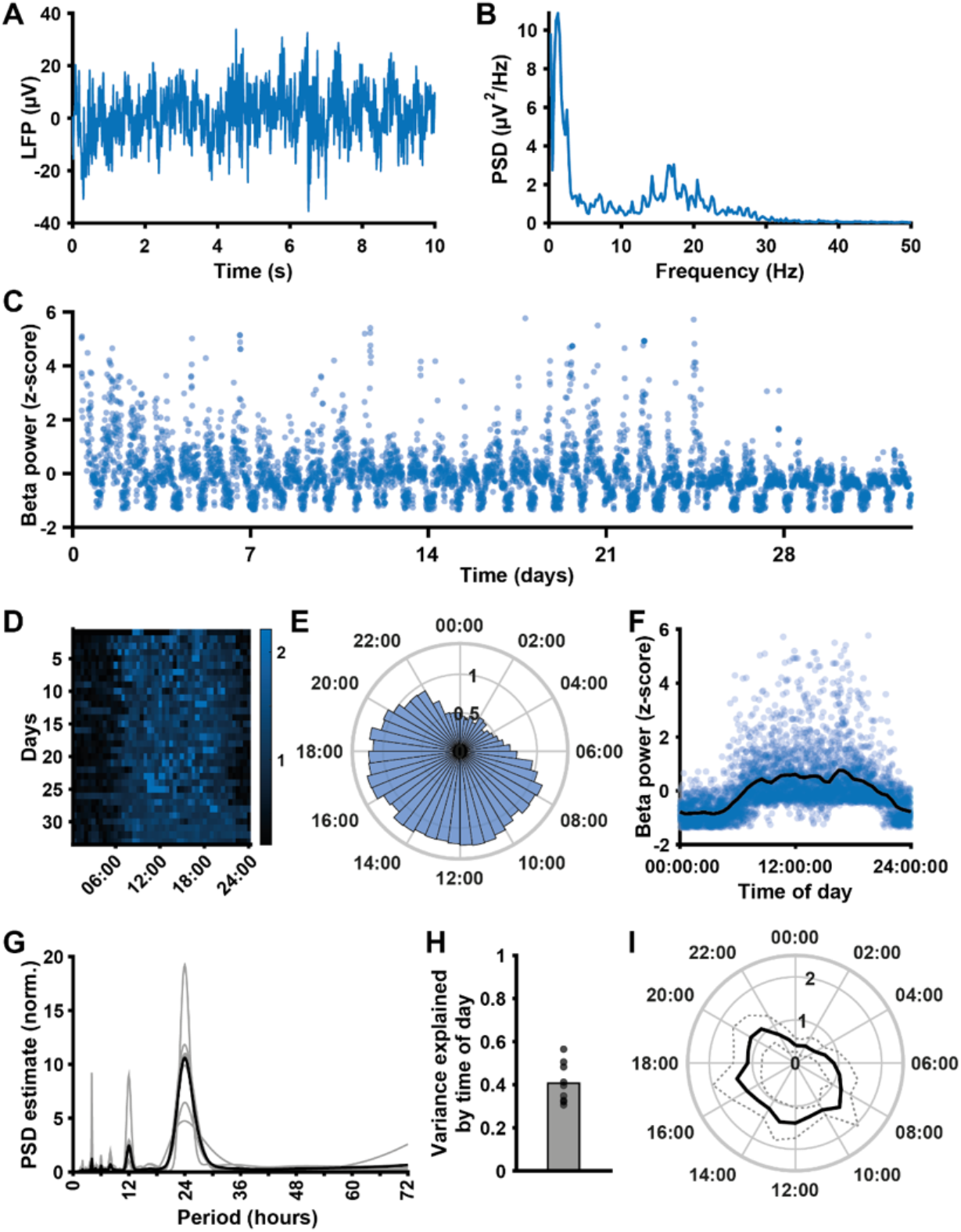
Beta oscillatory power measured with the Percept DBS device shows a consistent diurnal pattern with decreased beta during the night. **A:** A 10-second LFP recording from the STN of a Parkinson’s disease patient implanted with the Medtronic Percept DBS device, acquired during the BrainSense signal test. **B:** Power spectrum of the LFP segment in **A** (obtained using Welch’s method), showing a peak in the beta frequency range. **C:** Mean STN peak-to-peak beta power (µVp, sampled every 10 minutes) over a 1-month period, from the same example STN (after outlier removal). **D:** Heat map of beta power (detrended by normalising each day to its median value) across the 24 hours of the day for all days in the data collection period, for the same example STN. **E:** Detrended beta power across the 24-hour diurnal cycle generated from the data in **D**. For each day, the median beta power was calculated for each 30-minute time bin, and bar height in the circular bar graph represents the median across days. **F:** Normalised beta power measurements plotted against the time of day the measurements were taken for the same example STN, showing a consistent daily pattern of beta power. The black line represents a linear fit through the mean beta power for time of day (means obtained by dividing the day into 30-minute time bins). **G:** Periodogram of the beta power signal for all STN (normalised; estimated using Welch’s method). The population mean (black) highlights a clear peak at a period of 24h, which is present in all individual beta power time series (grey lines, n=9). **H:** The proportion of variance explained by time of day, estimated using fits such as illustrated in **D**, for all STN in the data set (mean 0.41±0.092; n=9; p<0.001 for all time series, temporal shuffling test). **I:** Mean (solid line) and standard deviation (dashed line) of detrended beta power around the diurnal cycle across all STN time series (n=9).

### Transitions between sleeping and waking periods are associated with beta fluctuations

To examine whether the diurnal profile of beta power observed was driven by periods of sleeping and waking, we acquired additional bilateral STN data from one patient (#5), who was asked to log their bedtime and rise time each day for a total period of 21 days (Figure 2A). Clear transitions in beta power were present around the times of going to bed and waking up of this patient (Figure 2A-C), and during estimated sleeping hours beta power was consistently low (Figure 2D). Consistent with this, there was a sharp increase in beta power slightly before their first scheduled medication time across patients (Figure 2E), and compared to the variance explained by time of day across the full diurnal cycle, time of day accounted for much less variance during day- or night-only epochs (Figure 2F; full 24h: 0.41±0.092; day only 0.13±0.11, p<0.001 vs 24h; night only 0.14±0.13, p=0.003 vs 24h; n=9 STN time series).

**Figure 2:**
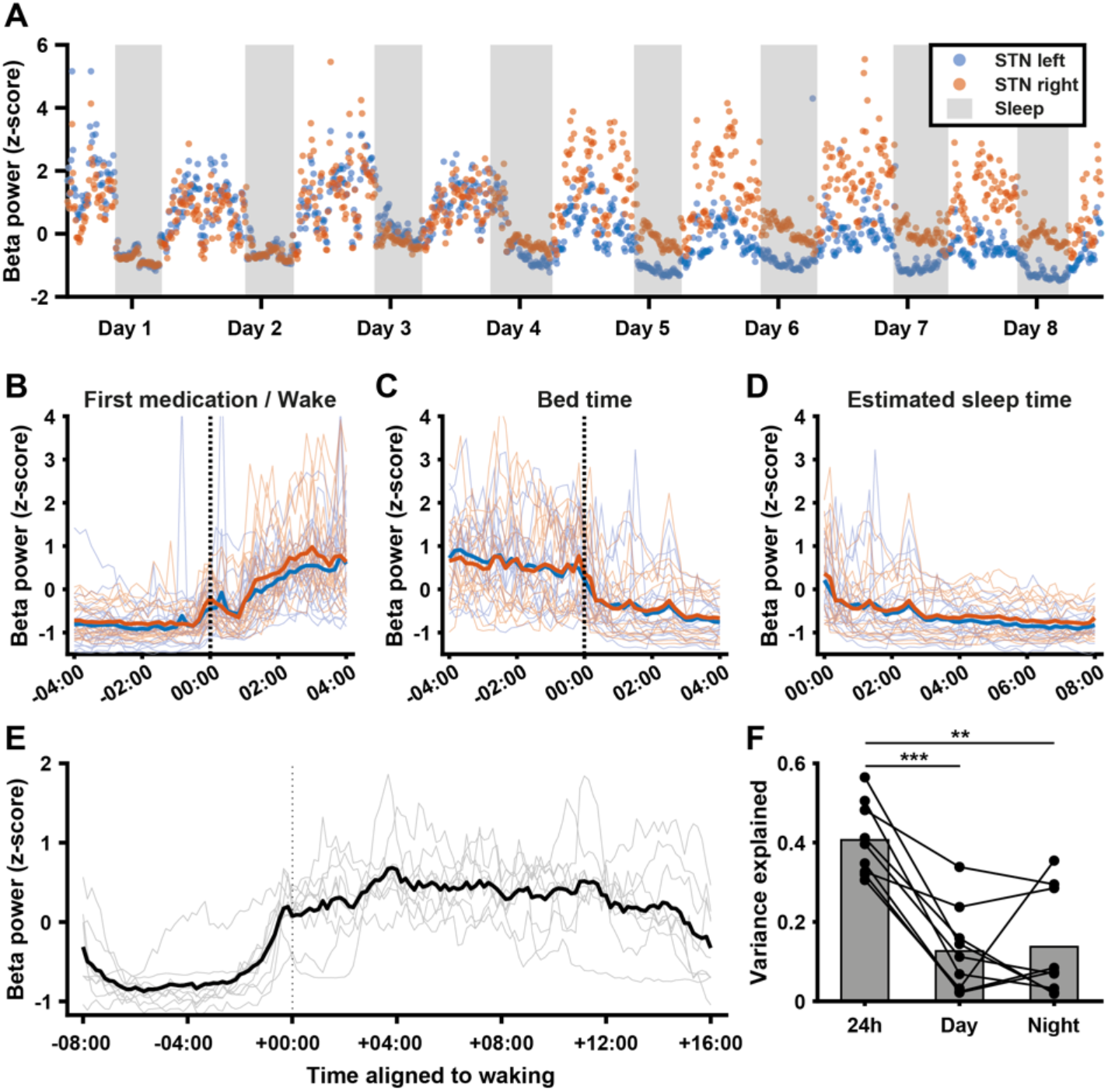
Diurnal variations in beta power are largely accounted for by transitions between sleeping and waking periods. **A:** Normalised beta power in the left and right STN (blue and orange, respectively) of one patient over a period of 8 days during which the patient recorded when they went to bed and when they rose in the morning. Grey areas represent time the patient was in bed for the night. **B**: Normalised beta power as in **A** for the full 21-day recording period, aligned to the time of waking. Blue and orange lines represent the mean for the left and right STN respectively, while thin blue and orange lines represent the corresponding individual time series. **C:** Normalised beta power as in **A** for the full 21-day recording period, aligned to the time of going to bed. Blue and orange thick lines represent the mean for the left and right STN respectively, while thin blue and orange lines represent the corresponding individual time series. **D:** Normalised beta power as in **A** for the full 21-day recording period, between going to bed and waking up. Blue and orange thick lines represent the mean for the left and right STN respectively, while thin blue and orange lines represent the corresponding individual time series. **E:** Average beta power aligned to estimated average wake up time for all STN time series in the data set (grey lines, n=9) and mean across all time series (thick black line). **F:** Variance explained by time of day across the whole 24h cycle vs. during the day or night alone. Full 24h: 0.41±0.092; Day only: 0.13±0.11, p<0.001 vs 24h; Night only: 0.14±0.13, p=0.003 vs. 24h; n=9 STN time series.

### Frequency-specificity of diurnal LFP power modulation

As the chronically streamed data represents the mean power in single frequency band, broadband fluctuations in LFP power are indistinguishable from fluctuations specific to the target frequency. To investigate whether the diurnal fluctuations we observed were specific to the beta band, we compared concurrently collected beta and contralateral theta power in 6 patients. If diurnal changes are driven by broad-band artifacts affecting both hemispheres, these signals should be highly correlated. Figure 3A&D show beta and theta band power for 2 patients (plotted against each other in Figure 3B&E), illustrating that beta and contralateral theta power could be either positively or negatively correlated. Overall, correlation coefficients between beta and theta power ranged from −0.27 to 0.61 (mean 0.13±0.34, n=6). Beta and theta power could also show different diurnal profiles (Figure 3C&F). In the beta signal, the proportion of variance explained by time of day was 0.34±0.24, while for the theta signal this was 0.21±0.10. While this difference was not significant (p=0.15), it demonstrates for individual cases that the diurnal patterns cannot be fully explained by broadband signal power. Indeed, across the patient population, beta band power was less uniformly distributed around the diurnal cycle than theta band power (Figure 3I&J).

**Figure 3.**
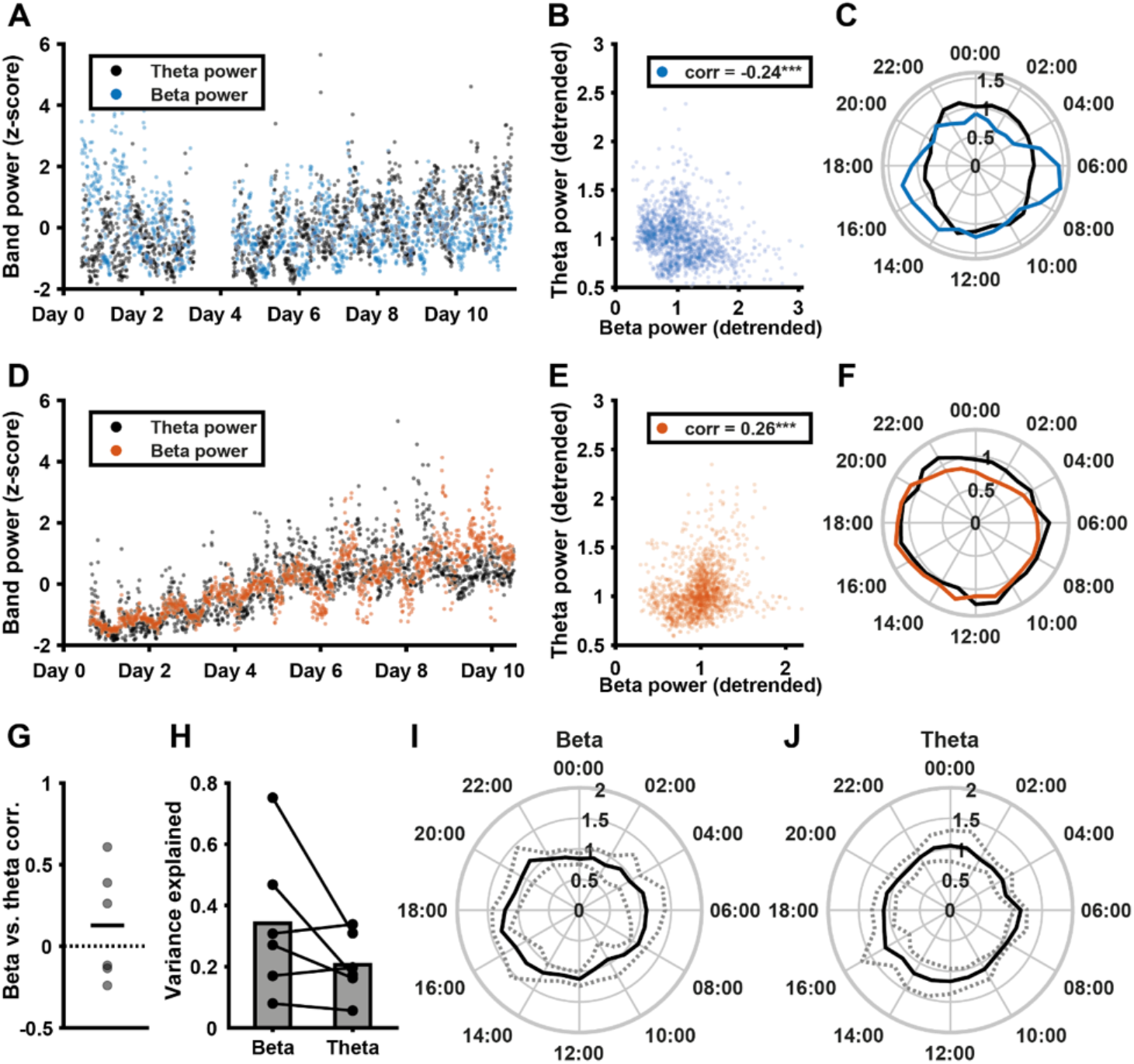
Correlations in long-term measurements of neural oscillatory power across frequency bands. **A**: Normalised beta power (blue) and theta power (black) collected concurrently from the left and right STN of one example patient. **B**: Scatter plot of detrended beta power vs detrended theta power shown in **A** (data was detrended by normalising values for each day to that day’s mean). For this patient, there was a negative correlation between beta and theta (r=-0.24, p<0.001). **C**: Median detrended beta (blue) and theta (black) power across the 24-hour diurnal cycle, showing different diurnal profiles of theta and beta in this example patient. **D**: Normalised beta power (orange) and theta power (black) collected concurrently from the left and right STN of a second example patient. **E**: Scatter plot of detrended beta power vs detrended theta power shown in **D** (data was detrended by normalising values for each day to that day’s mean). For this patient, there was a positive correlation between beta and theta (r=0.26, p<0.001). **F**: Median beta (orange) and theta (black) power across the 24-hour diurnal cycle, showing overlap in the diurnal profiles of beta and theta power in this second example patient. **G**: Pearson’s correlation coefficients for concurrently collected contralateral STN beta and theta power measurements (detrended; 0.13±0.34, n=6). **H**: Variance explained by time of day in concurrently collected beta and theta power measurements (p=0.15, n=6). **I**,**J**: Mean (solid black) and standard deviation (dashed, grey) concurrent diurnal beta (**I**) and theta (**J**) power profiles around the diurnal cycle, across the STN of 6 patients.

### Artifacts generated by everyday movements influence measures of LFP band power acquired with the Percept DBS device

The presence of correlations across frequency bands in several patients warranted further investigation what might underlie diurnal broad-band influences on the beta power measure. Movement artifacts could present a confound, as they would affect a large part of the power spectrum and would be more numerous during waking hours compared to during the night. We therefore asked a subset of patients to carry out a range of movements in the clinic while acquiring a bilateral STN LFP stream. Individual movements could have a clear impact on the LFP and the associated power spectrum (Figure 4A&B), and many movements increased both beta and theta power estimates (Figure 4C, Table 2). Particularly notable was a consistent effect of common head movements for all patients. Theta band power was increased even more than beta during most movements, with average increases of up to an order of magnitude.

**Table 2.** Common movements and their impact on measured beta and theta power (n=8 STN LFPs from 4 patients)

| Activity | Beta power ($\mu\text{Vp}$) | Relative beta (%) | Theta power ($\mu\text{Vp}$) | Relative theta (%) |
| --- | --- | --- | --- | --- |
| Rest | $3.76 \pm 3.82$ | $100 \pm 0$ | $3.31 \pm 2.20$ | $100 \pm 0$ |
| Walk | $3.39 \pm 2.11$ | $140 \pm 107$ | $8.18 \pm 8.33$ | $334 \pm 497$ |
| Turn | $4.45 \pm 2.49$ | $210 \pm 189$ | $8.04 \pm 6.89$ | $308 \pm 278$ |
| Stand | $3.08 \pm 2.42$ | $98 \pm 46$ | $3.88 \pm 3.17$ | $114 \pm 52$ |
| Sit | $3.81 \pm 3.17$ | $112 \pm 48$ | $4.57 \pm 3.77$ | $133 \pm 63$ |
| Stand up | $6.90 \pm 8.79$ | $427 \pm 858$ | $25.37 \pm 36.89$ | $1976 \pm 4595$ |
| Sit down | $3.17 \pm 2.35$ | $114 \pm 48$ | $6.61 \pm 5.08$ | $204 \pm 98$ |
| Arm swing | $3.67 \pm 2.35$ | $133 \pm 88$ | $5.91 \pm 4.00$ | $196 \pm 117$ |
| Arm lift | $4.81 \pm 4.05$ | $143 \pm 84$ | $17.73 \pm 29.24$ | $452 \pm 512$ |
| Arms to chest | $3.91 \pm 2.79$ | $131 \pm 67$ | $6.83 \pm 6.49$ | $192 \pm 88$ |
| Trunk bend | $2.89 \pm 1.78$ | $105 \pm 50$ | $5.94 \pm 5.01$ | $165 \pm 59$ |
| Trunk rotate | $2.59 \pm 1.68$ | $98 \pm 53$ | $5.05 \pm 4.97$ | $132 \pm 67$ |
| Head nod | $7.51 \pm 9.75$ | $172 \pm 55$ | $13.35 \pm 12.14$ | $433 \pm 460$ |
| Head shake | $7.52 \pm 7.92$ | $196 \pm 61$ | $22.18 \pm 24.41$ | $676 \pm 600$ |
| Head tilt | $15.53 \pm 29.83$ | $255 \pm 219$ | $60.67 \pm 150.2$ | $1218 \pm 2779$ |

**Figure 4:**
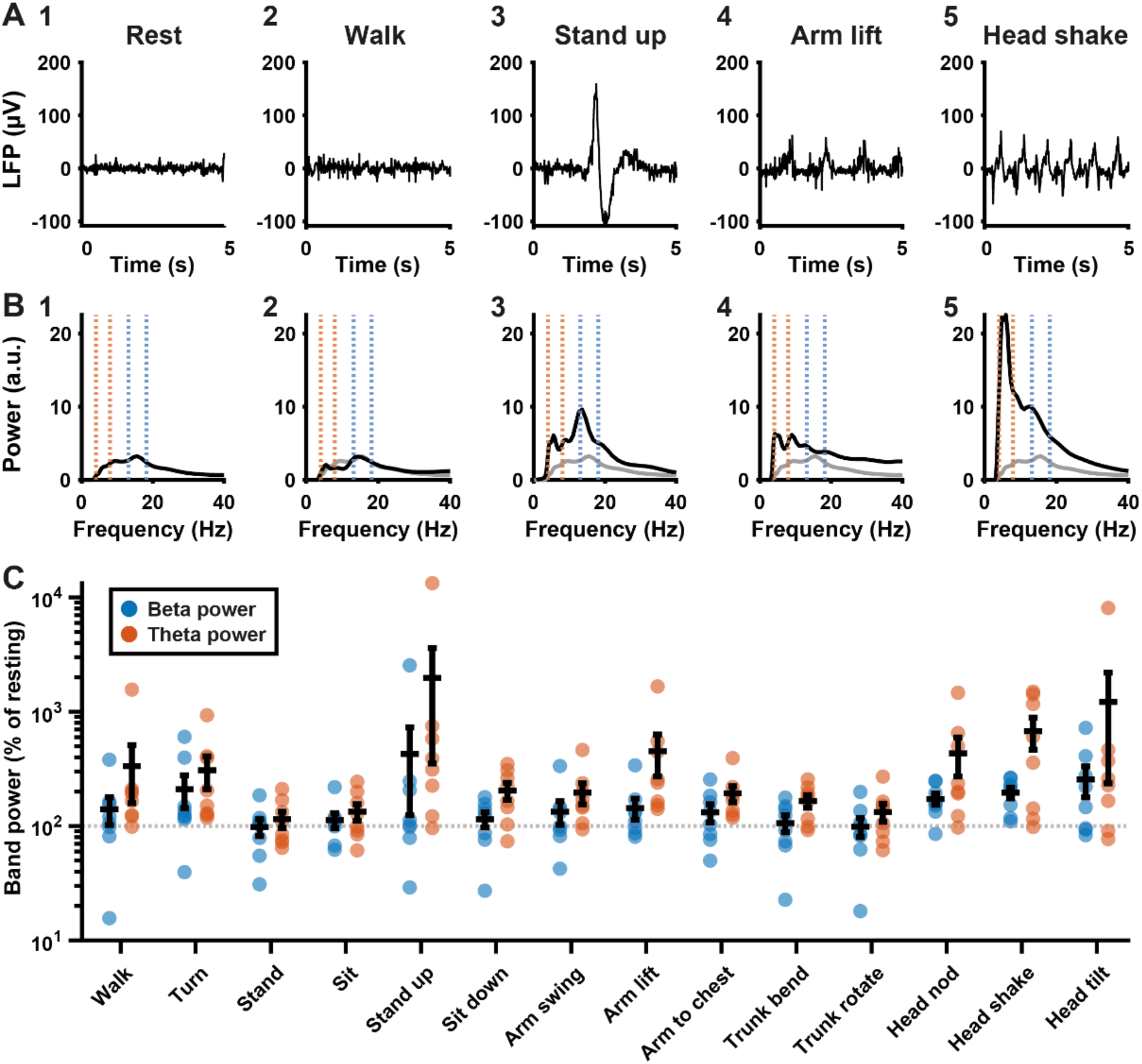
Common movements and their impact on measures of beta and theta band power. **A1-5:** 5-second LFP recordings from the STN of one patient collected with DBS on while they were resting (**A1**), walking (**A2**), standing up from a sitting position (**A3**), making a winging motion with the arms (**A4**) or shaking the head (**A5**) after application of a low pass filter at <98Hz and high pass filter at >4Hz. **B1-5:** Global wavelet power spectra (arbitrary units) corresponding to the LFP collected during the events depicted in **A1-5**. Blue dotted lines indicate the beta peak window for this STN, orange dotted lines indicate the theta band. The resting spectrum from **B1** is replicated as a grey line across the other spectra for comparison. **C:** Beta (blue) and theta (orange) band power measures as a percentage of resting power, from LFPs collected during a series of different movements (see Table 2 for all absolute and relative values; n=8 STN LFPs from 4 patients). The beta band was defined as 5Hz around the STN-specific beta peak, while the theta band was 4-8Hz for all patients.

To illustrate to what extent long-term data might be affected by movement artifacts, we collected in-clinic daytime measures on and off anti-parkinsonian medication and on and off DBS for two STN leads with different diurnal LFP profiles (Figure 5A1 and 5A2). One STN showed a smooth diurnal profile across the 24-hour cycle (5A1) while the other STN showed extreme intra-day peaks (5A2). This second patient had strong medication-related dyskinesia resulting in vigorous involuntary movement at certain times during the day. For the first STN, the measure collected in-clinic (on medication and with DBS) lies in the centre of the distribution of day-time long-term beta measurements, and to the right of most night-time measurements (Figure 5B1&C1). However, for the second STN, the in-clinic measure was much lower than most of the long-term daytime and even many night-time measurements. Moreover, the long-term data for daytime has a strongly bimodal distribution (Figure 5B2&5C2), consistent with the presence of dyskinesia-related movement artifacts. Day- and night-time distributions of beta power are provided for all patients in Appendix 1, Supplementary Figure S1-6, G1&2.

**Figure 5:**
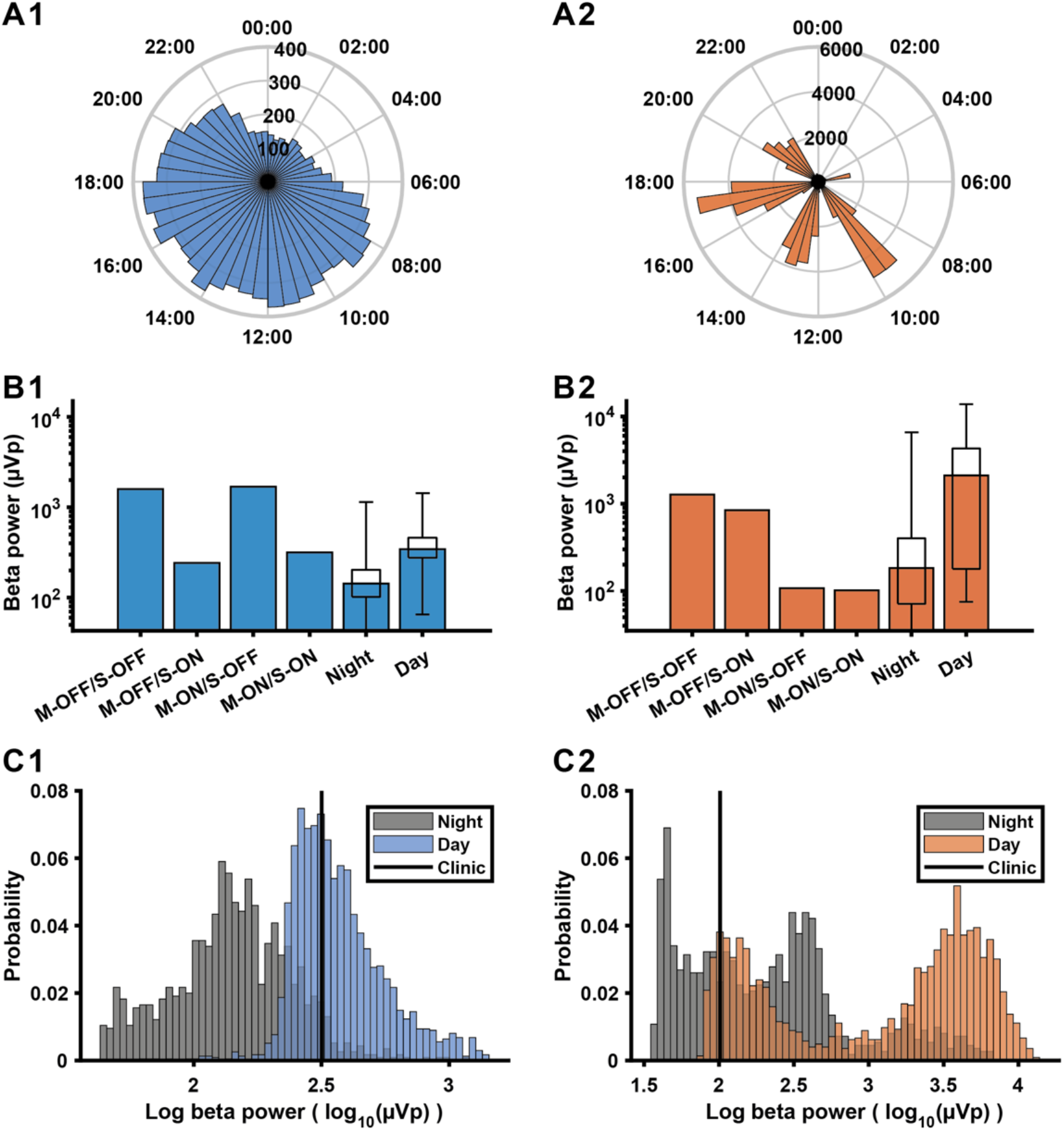
A comparison of beta power measures collected in-clinic and during long-term data collection. **A1**,**2:** Diurnal profile of beta power across the 2h cycle from 2 STN leads from different patients. Each bar represents the median beta power (µVp) in the time bin across all days. **B1**,**2:** In-clinic measures of beta power collected with DBS on and off (S-ON/S-OFF), while on and off medication (M-ON/M-OFF), compared to the median and distribution of beta power values (boxplots) obtained from long-term data collection during night-time (00:00-06:00) and daytime (08:00-20:00), for the same two examples. **C1**,**2:** Distributions of daytime (08:00-20:00) and night-time (00:00-06:00) log-transformed beta power measurements (log_10_(µVp)) during the long-term sensing period for the same examples. The black line indicates the in-clinic measurement taken while on dopaminergic medication during DBS on.

## Discussion

Using a chronically sensing DBS device, we show that the amplitude of beta oscillations during continuous DBS follows a consistent diurnal pattern. We demonstrate for the first time that this can be characterised using a commercially available device used for the treatment of PD. While these findings offer new avenues for adaptive control of DBS, we also report that common movements can confound the beta power measure. Our approach highlights the value of taking a longer-timescale perspective on data from DBS devices and has important implications for the future implementation of aDBS.

### Physiological basis of diurnal fluctuations in beta power during continuous DBS

We found that STN beta power was consistently higher during the day, and lower during the night, across a group of patients undergoing continuous high-frequency DBS. The pattern of modulation closely followed the estimated sleep schedule of patients. This may be explained by changes in the physiological mechanisms underlying the generation of beta oscillations during slow wave sleep. Whole-night recordings of STN-LFPs show that the beta frequency activity is reduced by around half in NREM sleep stages compared to REM and wakefulness (Thompson et al., 2018). In contrast, slow and delta frequencies increase, in line with the key role of these frequencies in thalamocortical sleep structure (Steriade et al., 1993). In parkinsonian rodents recorded under anaesthesia, the emergence of beta oscillations across the cortex, basal ganglia and thalamus is dependent on the absence of slow wave activity (Mallet et al., 2008; Nakamura et al., 2021; Sharott et al., 2017). It should be noted, however, that during NREM sleep in parkinsonian primates, beta oscillations are interspersed with slow oscillations and may underlie sleep disturbance in PD (Mizrahi-Kliger et al., 2020). In our study, all beta power measurements were conducted during continuous DBS, which has previously been shown to improve sleep quality (Baumann-Vogel et al., 2017). Recordings of wide-band signals over multiple nights, together with detailed polysomnography and sleep diaries, will likely be needed to fully understand how beta, NREM activities and sleep quality are related in PD.

### Influence of artifacts on the Percept long term data

We have recently highlighted the propensity of ECG artifacts to corrupt signals recorded from devices with chronic sensing capability (Neumann et al., 2021). Heart rate is modulated by the brain’s internal clock, slowing during the night and gradually rising to a peak in the afternoon (Black et al., 2019), and could thus contribute to the diurnal fluctuations seen here. By using the Perceive toolbox to screen our data, we were able to exclude ECG as a major contributing factor here. In contrast, delineating the influence of movement artifacts on chronic sensing data is more challenging. We showed that LFP streams from the Percept DBS implant can be heavily affected by common movements. Head movements, in particular, yielded band power estimates above resting levels across patients. To address the influence of these artifacts, we interrogated the interdependence of theta and beta amplitude across hemispheres. If broad-band movement or ECG artifacts were the primary driver of diurnal modulation in both hemispheres, theta and beta should have been highly correlated and theta would be similarly or more strongly locked to time of day. However, in most patients the correlation between bands was either weakly positive or negative, and beta power, on average, showed a stronger influence of time of day. Moreover, our characterisation of beta amplitude during movement was made over short LFP segments (the seconds surrounding the movement of interest) while long-term sensing with the Percept generates a measure of average power over much longer epochs (10 minutes). Therefore, the impact of short, isolated movements on the long-term sensing data may be modest in most patients. Nevertheless, the streamed signal is vulnerable to influence from such movements if they are repeated or sustained during a large proportion of a 10-minute epoch. This is highlighted in our recordings from a patient with regular episodes of dyskinesia (see Figure 5). Overall, it is likely that, in most patients, diurnal fluctuations in the streamed signal reflect physiological changes in LFP beta power. However, if artifacts associated with specific movements occur frequently enough and cluster together, they may significantly confound the signal.

### Implications for adaptive deep brain stimulation

There are currently two main approaches to adaptive stimulation for PD. The first is to deliver high-frequency stimulation only when the amplitude is above a set threshold, resulting in modulation of stimulation at a sub-second timescale equivalent to that of beta bursts (Little et al., 2013). An alternative method is to adjust the amplitude of continuous 130Hz stimulation proportional to fluctuations in the beta amplitude on the scale of minutes (Rosa et al., 2015). Our recordings suggest beta amplitude consistently drops by around 50% during the night. In either aDBS strategy, the result would be that the amount/amplitude of stimulation is significantly reduced at night. This strategy is not consistent with the preference of PD patients, who report increased bradykinesia during wake-up periods at night if stimulation is withdrawn (Lyons & Pahwa, 2006). Taken together with our findings, beta amplitude, DBS occurrence/amplitude and clinical outcome appear to have a different relationship during the day and night, which may require customised control.

In the “desired” situation, where the input to the control loop is driven by physiological changes in beta amplitude (Figure 6A), we suggest three possible strategies for dealing with the reduction in beta amplitude during sleep. The first method is to simply revert to open loop stimulation during the time epoch when a patient is expected to be sleeping (Gilron et al., 2021), which can be achieved with simple clock-based control. The second method is to make the threshold for the classifier a function of time. The variable threshold will help compensate for the deterministic time-dependent variance of the beta amplitude. The trade-off with this approach is the potential difficulty in setting an optimal value, but emerging technologies such as the loop recording of the Percept can help to gather this information. The third method would provide more nuanced adjustments based on the immediate sleep stage, where the beta band might cycle through the normal progression from NREM to REM sleep. All these methods include a feedforward, time-based adjustment to the feedback pathway (Zamora et al., 2021). In practice, the optimal approach will be a balance of patient benefit versus clinical setup burden.

**Figure 6:**
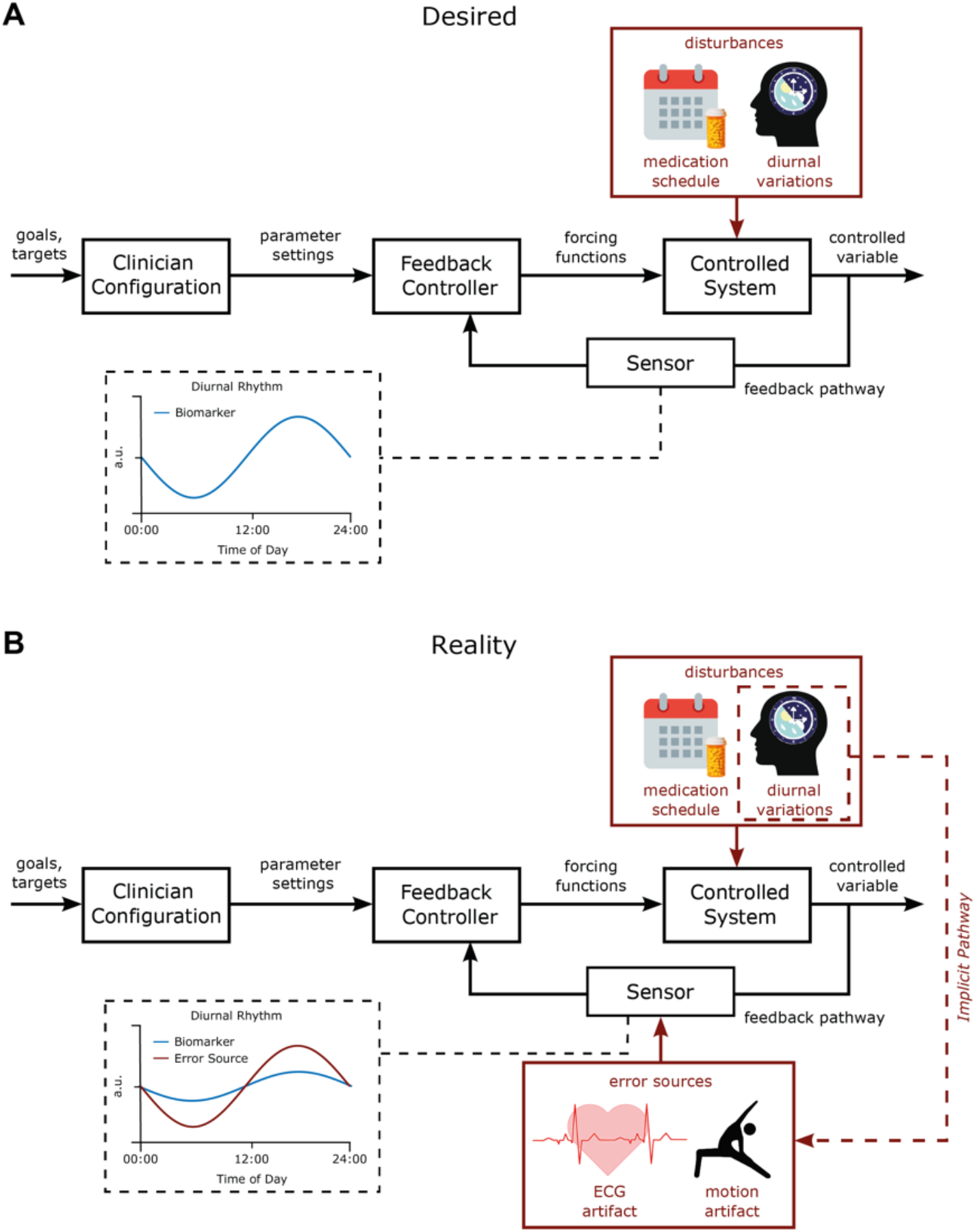
Desired vs. Realistic block diagram of chronoadaptive feedback DBS control system. **A:** Desired control system. In the desired DBS control system, therapeutic goals (i.e. % suppression of beta power) are specified by clinicians for specific times of the day. A feedback controller modulates the DBS parameters to maintain the controlled variable, the biomarker, at the specified target level. Time-based adjustments of control goals synchronized to patient-specific chronotypes can then be incorporated to accommodate known disturbances such as the medication-schedule and diurnal variation of the monitored biomarker. In this manner, time-localized adjustments to stimulation parameters are provided in response to physiological variations. **B:** Actual control system. In the actual control system at present diurnal variations in artifact sources may couple into the sensor in the feedback pathway. This can confound observations of physiological fluctuations in the monitored biomarker signal due to the amplitude of artefacts being greater than the signal of interest.

### Potential confounds to adaptive DBS from artifacts

Our results highlight that as patients go about their daily lives, they will behave in ways that have the potential to drive non-physiological fluctuations in the streamed amplitude data. Low-frequency oscillations, including the beta range, appear particularly vulnerable to ECG and movement artifacts. In terms of threshold-based adaptive stimulation, this could lead to a situation whereby the threshold for stimulation is set in the “desired” condition but is then inappropriately triggered in real-life conditions when artifacts are present (Figure 6B). In PD, movement artifacts that constantly push the signal above the trigger threshold would result in continuous stimulation. This would mimic conventional continuous DBS, and any potential benefits of adaptive stimulation (e.g. side effect control) would be lost, a situation that appears particularly likely to occur in patients with dyskinesia. Movement-based confounds could be more serious in applications when dual threshold aDBS is implemented with high upper stimulation amplitude, where movement artifacts could maintain a sub-optimally high level of stimulation. To further complicate these issues, physiological circadian or diurnal rhythms are implicitly synchronised to the timescale of ECG and movement artifacts through physical activity (Figure 6B). The contribution of these artifacts will differ based on the signal-to-noise ratio of the movement artefact and the physiological signal. For example, in some patients beta amplitude in sleep may be physiologically driven, but in wakefulness be a composite of physiology and artifact (Figure 6B).

As the use of devices with sensing capability develops, it will be crucial to evaluate the composite of physiological and artifactual influences on any input signal used either to investigate pathophysiological and/or to control stimulation. Pre-screening for artifact contamination from the ECG or movements is an important first step towards this. Ideally, access to the full-spectrum LFP signal underlying the continuous band power measurements would provide clinicians and researchers with a means to evaluate the integrity of the data. When using devices without this functionality, regular inspection of the distribution of values returned by the system may provide an indication of contamination, as in the dyskinetic patient described here. When sensing is used for aDBS, systems should be set up in such a way that transient or prolonged artifact influence cannot raise or lower overall stimulation levels to dangerous or poorly tolerated levels (Gunduz et al., 2019). Finally, on-line detection of extreme values or a suspicious measurement distribution could be set to engage a pre-determined fail-safe mode, reverting the system to a well-tolerated level of stable DBS. Development of cranially mounted devices may further help to avoid ECG and movement artefacts (Neumann et al., 2021; Sorkhabi et al., 2020).

## Conclusion

In conclusion, beta activity in PD shows significant diurnal fluctuations with a reduction at night that may decrease aDBS to suboptimal stimulation levels. Moreover, artifacts resulting from patient activities remain a concern for applications of chronic sensing. Nevertheless, chronic recordings of band-specific LFP amplitude provide the opportunity to define the chronotype of a disease-relevant activity. Access to this information provides many opportunities to improve therapy, both through better informed adjustments made by the clinician and through the development of feedforward and feedback strategies to improve adaptive DBS.

## Data Availability

Data produced in the present study are currently not yet available for use by those not directly involved in the research project

## Acknowledgements

Most of all, we would like to thank the patients who participated in our study. We also thank Ulrike Uhlig for help with patient management and recordings.

## Appendix 1: Supplementary patient overview figures

**Figure S1:**
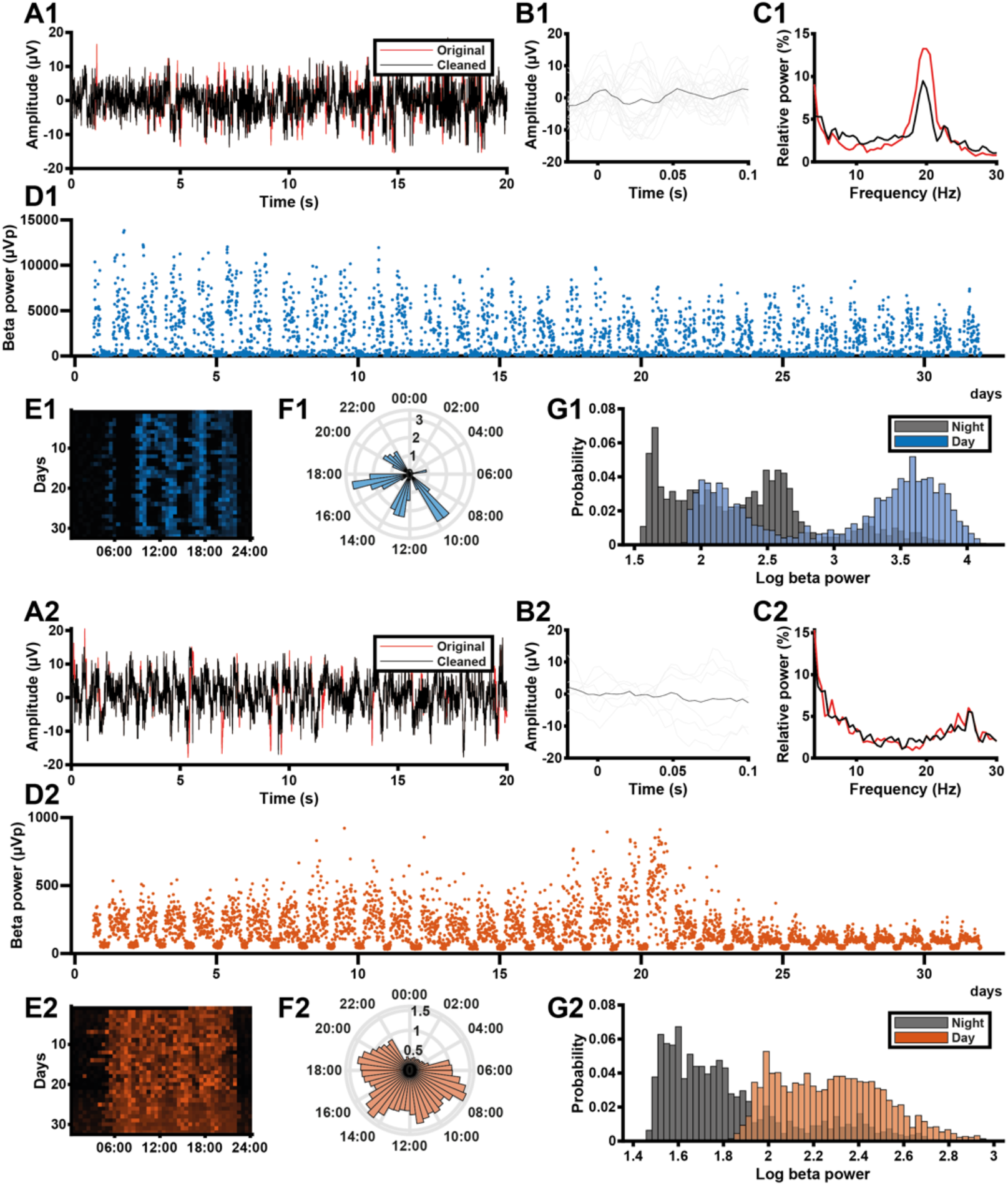
Patient #1 individual data overview. **A1**,**2**: LFP recorded from the left (1) and right (2) STN of a PD patient during the Percept’s BrainSense signal test. Red line represents the original signal, black represents the signal that has been recovered using the Perceive toolbox (Neumann et al. 2021). **B1**,**2**: ECG waveforms detected in the LFP signals in A using the Perceive toolbox. Grey lines represent individual candidate waveforms, the black line represents the average waveform estimate. **C1**,**2**: Normalised Welch’s power spectra of the original (red) and cleaned (black) LFP signals in A. **D1**,**2**: Beta power values (µVp) recorded from the left (1) and right (2) STN of this PD patient during the long-term data collection period (outliers with z-score > 6 removed). **E1**,**2**: Heat map of beta power (detrended by normalising each day to its median value) across the 24 hours of the day for all days in the data collection period, for the same example STN. **F1**,**2**: Detrended beta power across the 24-hour diurnal cycle generated from the data in **D**. For each day, the median beta power was calculated for each 30-minute time bin, and bar height in the circular bar graph represents the median across days. **G1**,**2**: Distributions of daytime (08:00-20:00) and night-time (00:00-06:00) beta power measurements in D, log-transformed (log_10_(µVp)).

**Figure S2:**
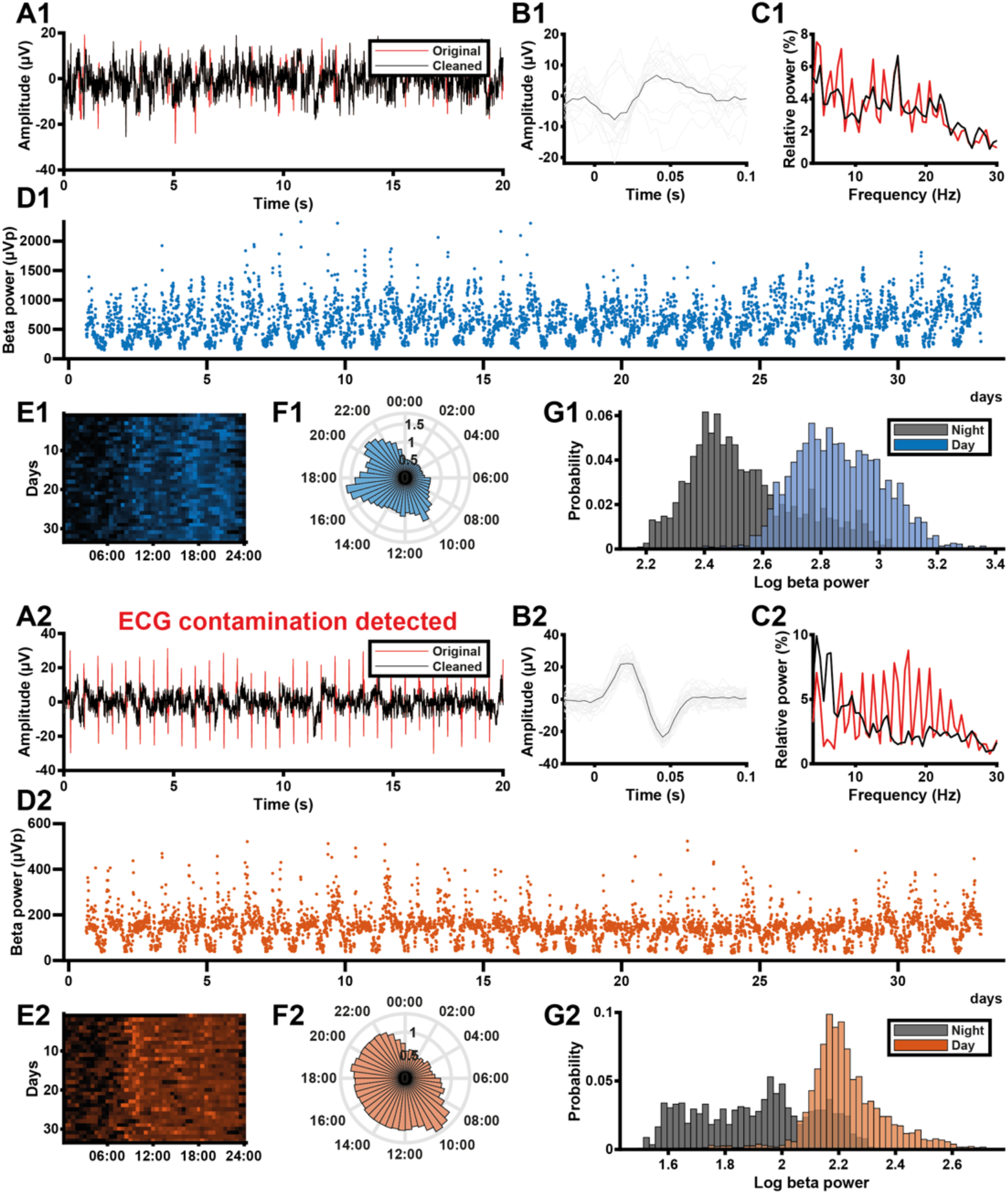
Patient #2 individual data overview. **A1**,**2**: LFP recorded from the left (1) and right (2) STN of a PD patient during the Percept’s BrainSense signal test. Red line represents the original signal, black represents the signal that has been recovered using the Perceive toolbox (Neumann et al. 2021). **B1**,**2**: ECG waveforms detected in the LFP signals in A using the Perceive toolbox. Grey lines represent individual candidate waveforms, the black line represents the average waveform estimate. **C1**,**2**: Normalised Welch’s power spectra of the original (red) and cleaned (black) LFP signals in A. **D1**,**2**: Beta power values (µVp) recorded from the left (1) and right (2) STN of this PD patient during the long-term data collection period (outliers with z-score > 6 removed). **E1**,**2**: Heat map of beta power (detrended by normalising each day to its median value) across the 24 hours of the day for all days in the data collection period, for the same example STN. **F1**,**2**: Detrended beta power across the 24-hour diurnal cycle generated from the data in **D**. For each day, the median beta power was calculated for each 30-minute time bin, and bar height in the circular bar graph represents the median across days. **G1**,**2**: Distributions of daytime (08:00-20:00) and night-time (00:00-06:00) beta power measurements in D, log-transformed (log_10_(µVp)).

**Figure S3:**
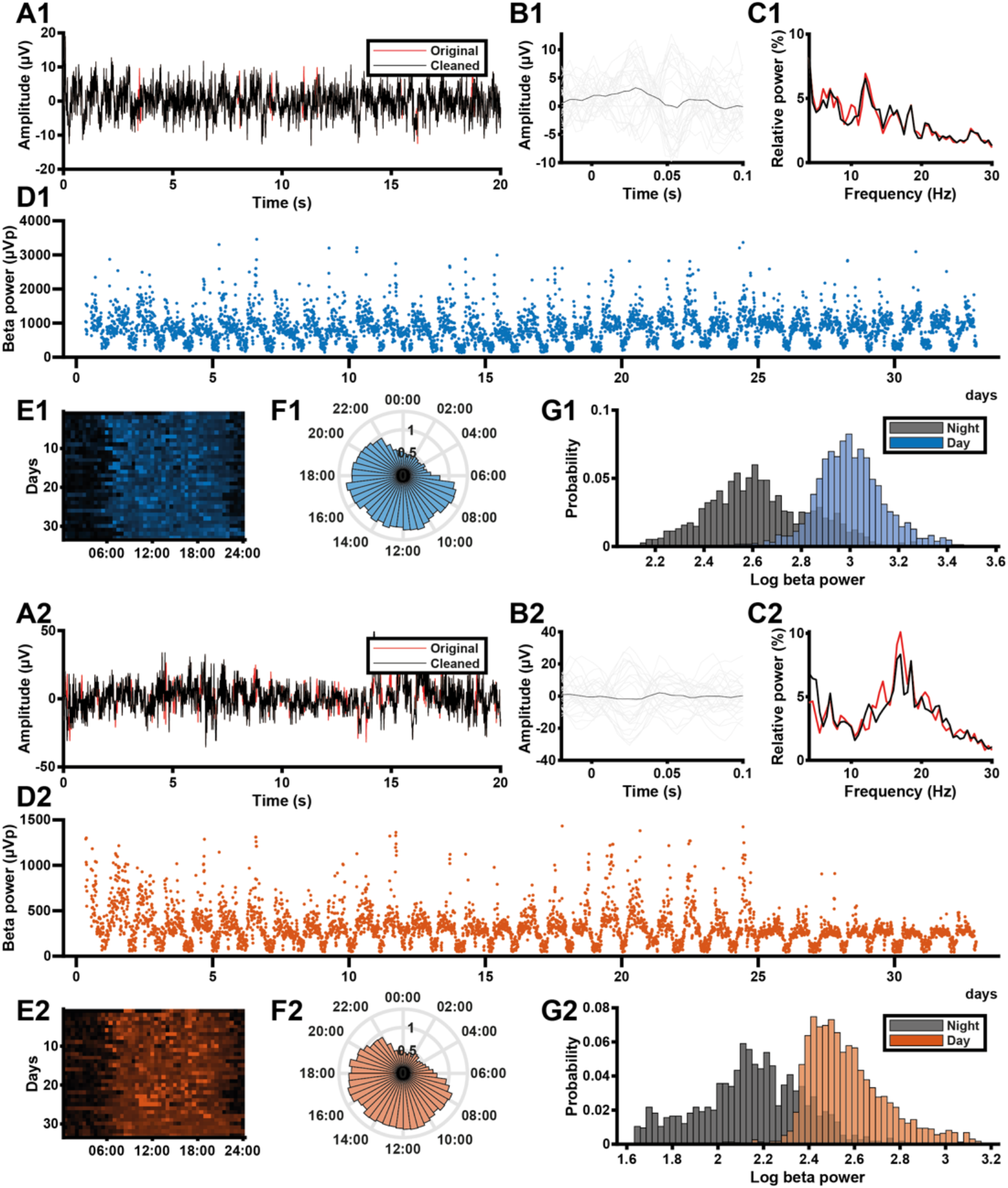
Patient #3 individual data overview. **A1**,**2**: LFP recorded from the left (1) and right (2) STN of a PD patient during the Percept’s BrainSense signal test. Red line represents the original signal, black represents the signal that has been recovered using the Perceive toolbox (Neumann et al. 2021). **B1**,**2**: ECG waveforms detected in the LFP signals in A using the Perceive toolbox. Grey lines represent individual candidate waveforms, the black line represents the average waveform estimate. **C1**,**2**: Normalised Welch’s power spectra of the original (red) and cleaned (black) LFP signals in A. **D1**,**2**: Beta power values (µVp) recorded from the left (1) and right (2) STN of this PD patient during the long-term data collection period (outliers with z-score > 6 removed). **E1**,**2**: Heat map of beta power (detrended by normalising each day to its median value) across the 24 hours of the day for all days in the data collection period, for the same example STN. **F1**,**2**: Detrended beta power across the 24-hour diurnal cycle generated from the data in **D**. For each day, the median beta power was calculated for each 30-minute time bin, and bar height in the circular bar graph represents the median across days. **G1**,**2**: Distributions of daytime (08:00-20:00) and night-time (00:00-06:00) beta power measurements in D, log-transformed (log_10_(µVp)).

**Figure S4:**
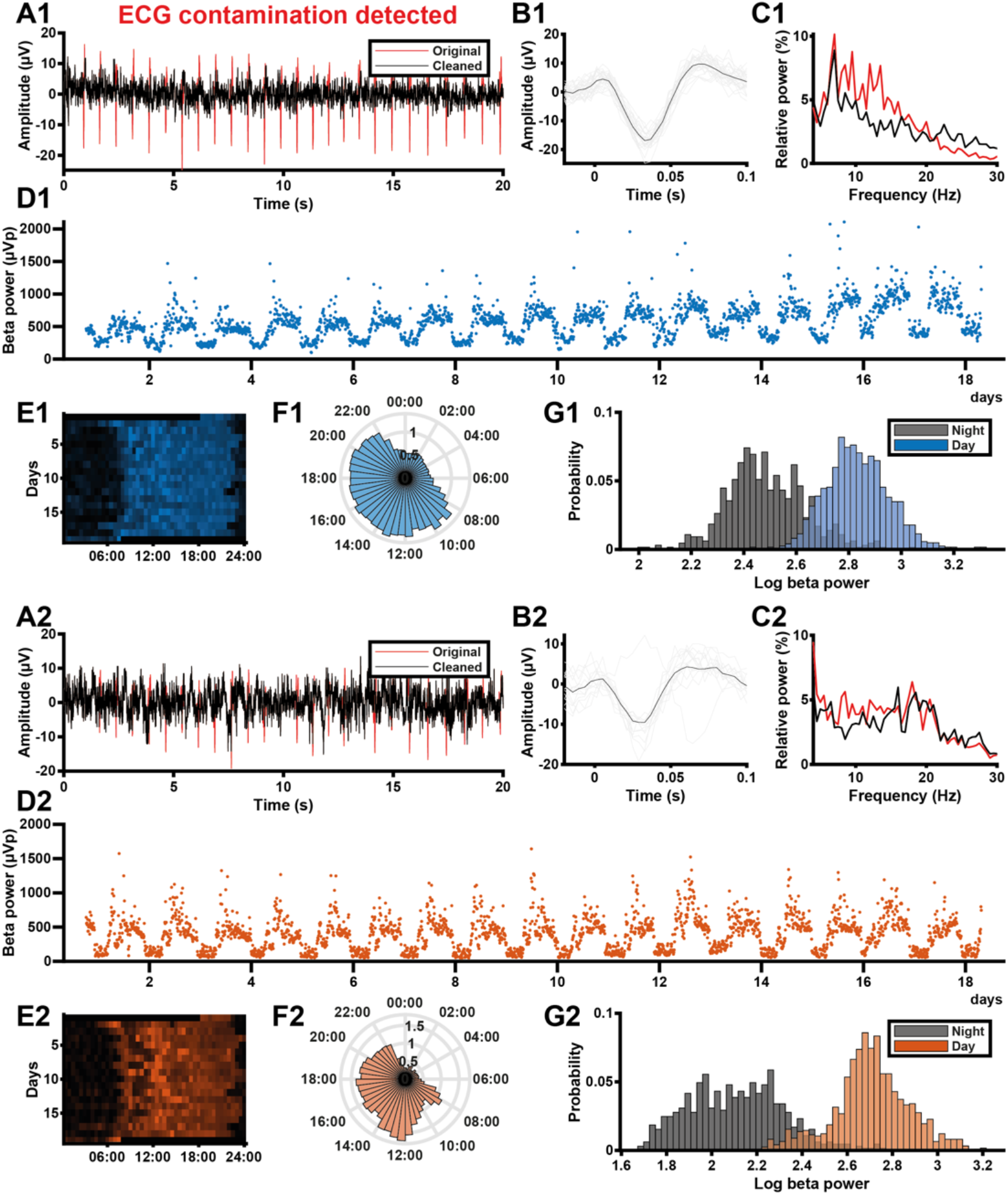
Patient #4 individual data overview. **A1**,**2**: LFP recorded from the left (1) and right (2) STN of a PD patient during the Percept’s BrainSense signal test. Red line represents the original signal, black represents the signal that has been recovered using the Perceive toolbox (Neumann et al. 2021). **B1**,**2**: ECG waveforms detected in the LFP signals in A using the Perceive toolbox. Grey lines represent individual candidate waveforms, the black line represents the average waveform estimate. **C1**,**2**: Normalised Welch’s power spectra of the original (red) and cleaned (black) LFP signals in A. **D1**,**2**: Beta power values (µVp) recorded from the left (1) and right (2) STN of this PD patient during the long-term data collection period (outliers with z-score > 6 removed). **E1**,**2**: Heat map of beta power (detrended by normalising each day to its median value) across the 24 hours of the day for all days in the data collection period, for the same example STN. **F1**,**2**: Detrended beta power across the 24-hour diurnal cycle generated from the data in **D**. For each day, the median beta power was calculated for each 30-minute time bin, and bar height in the circular bar graph represents the median across days. **G1**,**2**: Distributions of daytime (08:00-20:00) and night-time (00:00-06:00) beta power measurements in D, log-transformed (log_10_(µVp)).

**Figure S5:**
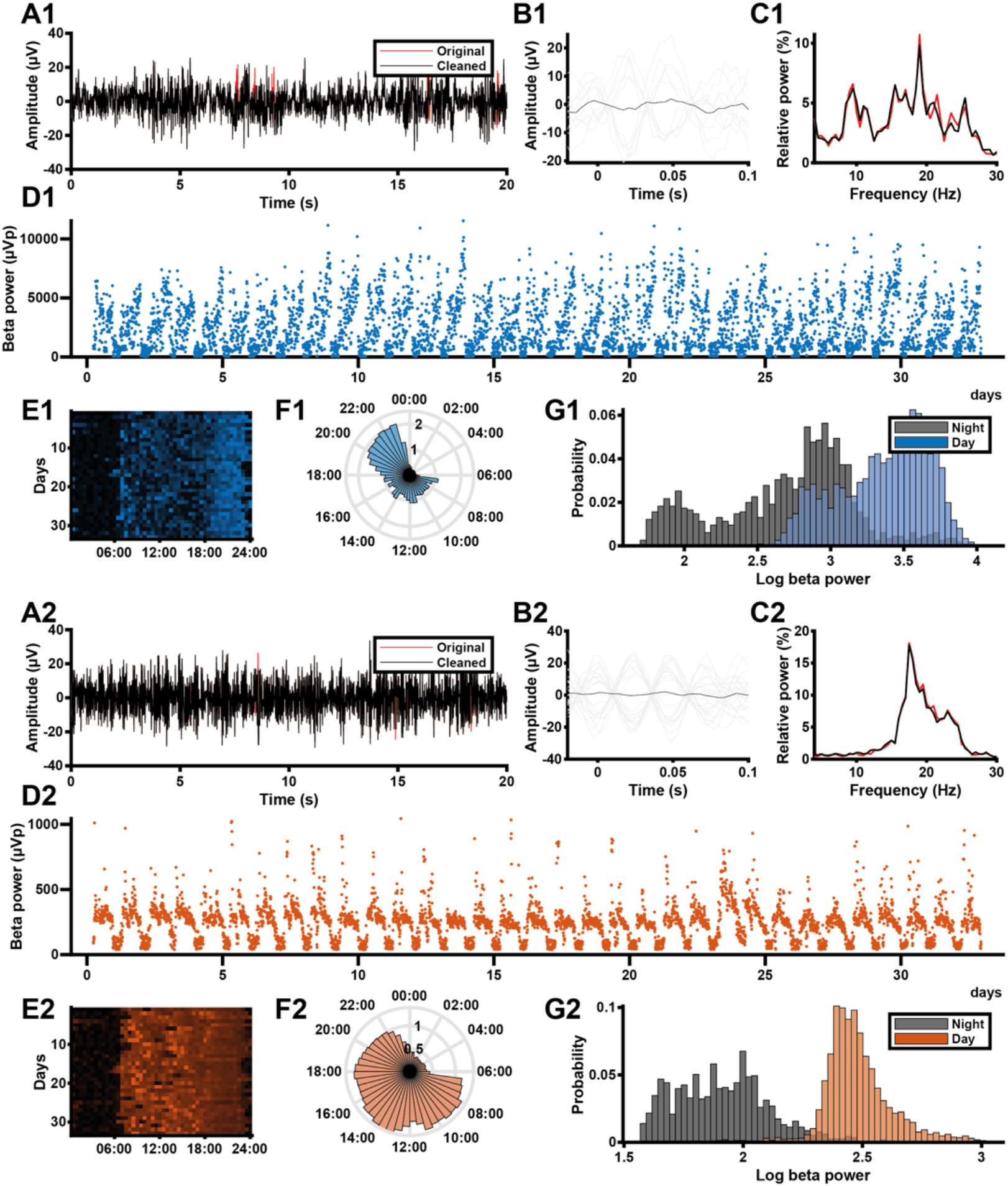
Patient #5 individual data overview. **A1**,**2**: LFP recorded from the left (1) and right (2) STN of a PD patient during the Percept’s BrainSense signal test. Red line represents the original signal, black represents the signal that has been recovered using the Perceive toolbox (Neumann et al. 2021). **B1**,**2**: ECG waveforms detected in the LFP signals in A using the Perceive toolbox. Grey lines represent individual candidate waveforms, the black line represents the average waveform estimate. **C1**,**2**: Normalised Welch’s power spectra of the original (red) and cleaned (black) LFP signals in A. **D1**,**2**: Beta power values (µVp) recorded from the left (1) and right (2) STN of this PD patient during the long-term data collection period (outliers with z-score > 6 removed). **E1**,**2**: Heat map of beta power (detrended by normalising each day to its median value) across the 24 hours of the day for all days in the data collection period, for the same example STN. **F1**,**2**: Detrended beta power across the 24-hour diurnal cycle generated from the data in **D**. For each day, the median beta power was calculated for each 30-minute time bin, and bar height in the circular bar graph represents the median across days. **G1**,**2**: Distributions of daytime (08:00-20:00) and night-time (00:00-06:00) beta power measurements in D, log-transformed (log_10_(µVp)).

**Figure S6:**
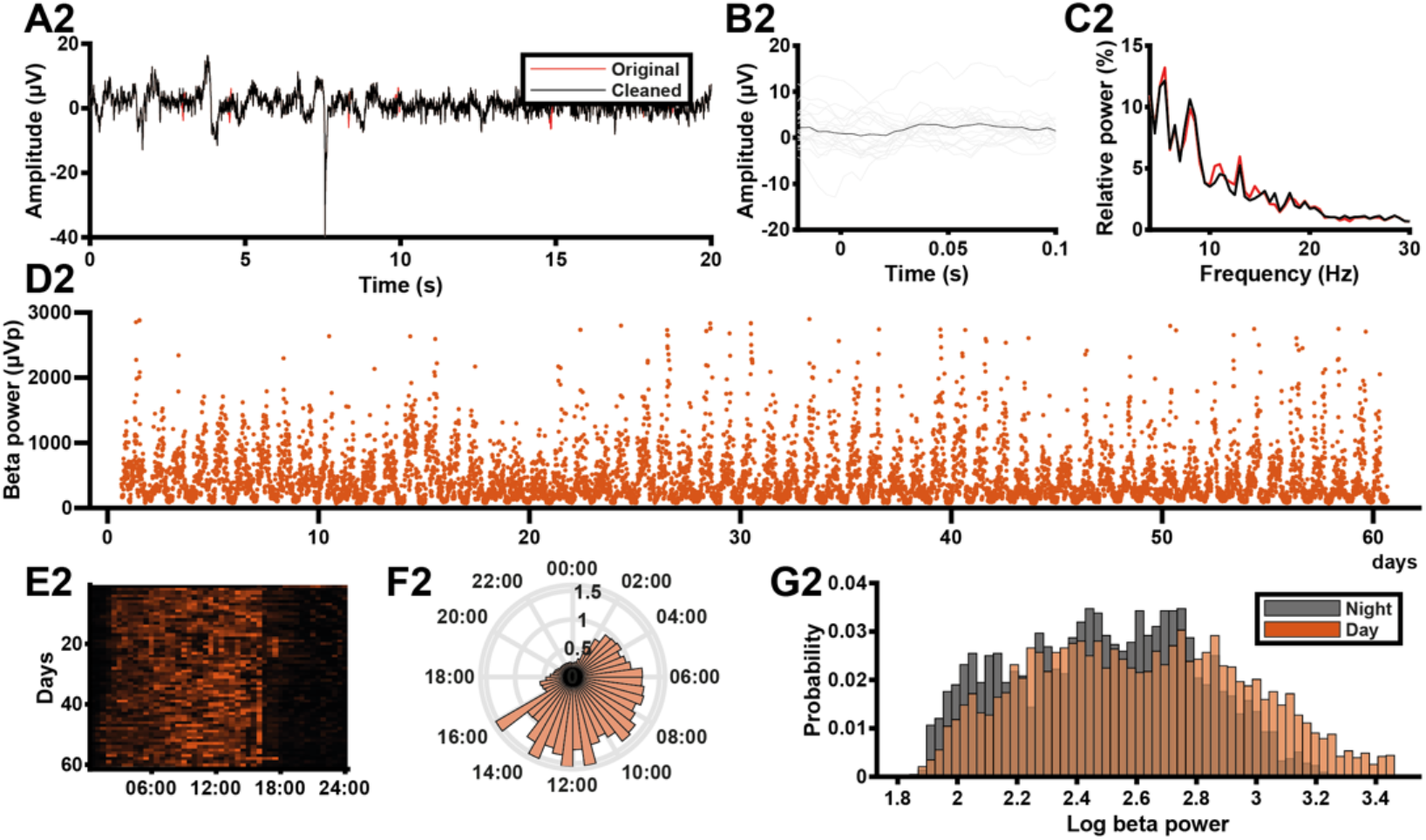
Patient #6 individual data overview (left side not available for this patient) **A1**,**2**: LFP recorded from the left (1) and right (2) STN of a PD patient during the Percept’s BrainSense signal test. Red line represents the original signal, black represents the signal that has been recovered using the Perceive toolbox (Neumann et al. 2021). **B1**,**2**: ECG waveforms detected in the LFP signals in A using the Perceive toolbox. Grey lines represent individual candidate waveforms, the black line represents the average waveform estimate. **C1**,**2**: Normalised Welch’s power spectra of the original (red) and cleaned (black) LFP signals in A. **D1**,**2**: Beta power values (µVp) recorded from the left (1) and right (2) STN of this PD patient during the long-term data collection period (outliers with z-score > 6 removed). **E1**,**2**: Heat map of beta power (detrended by normalising each day to its median value) across the 24 hours of the day for all days in the data collection period, for the same example STN. **F1**,**2**: Detrended beta power across the 24-hour diurnal cycle generated from the data in **D**. For each day, the median beta power was calculated for each 30-minute time bin, and bar height in the circular bar graph represents the median across days. **G1**,**2**: Distributions of daytime (08:00-20:00) and night-time (00:00-06:00) beta power measurements in D, log-transformed (log_10_(µVp)).

