## Supplementary figures and images for "Diurnal modulation of subthalamic beta oscillatory power in Parkinson’s disease patients during deep brain stimulation"

### Figure 1

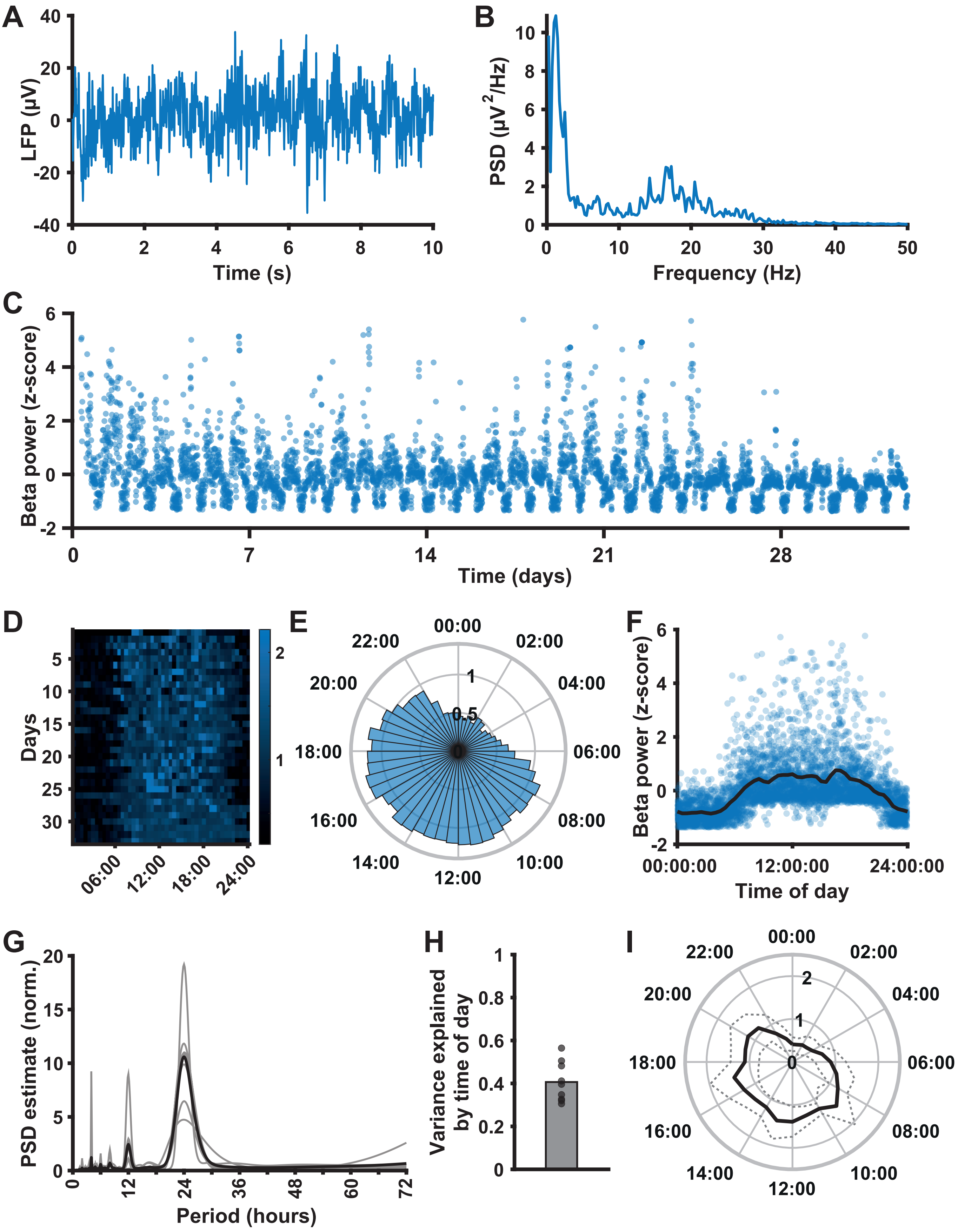

### Figure 2

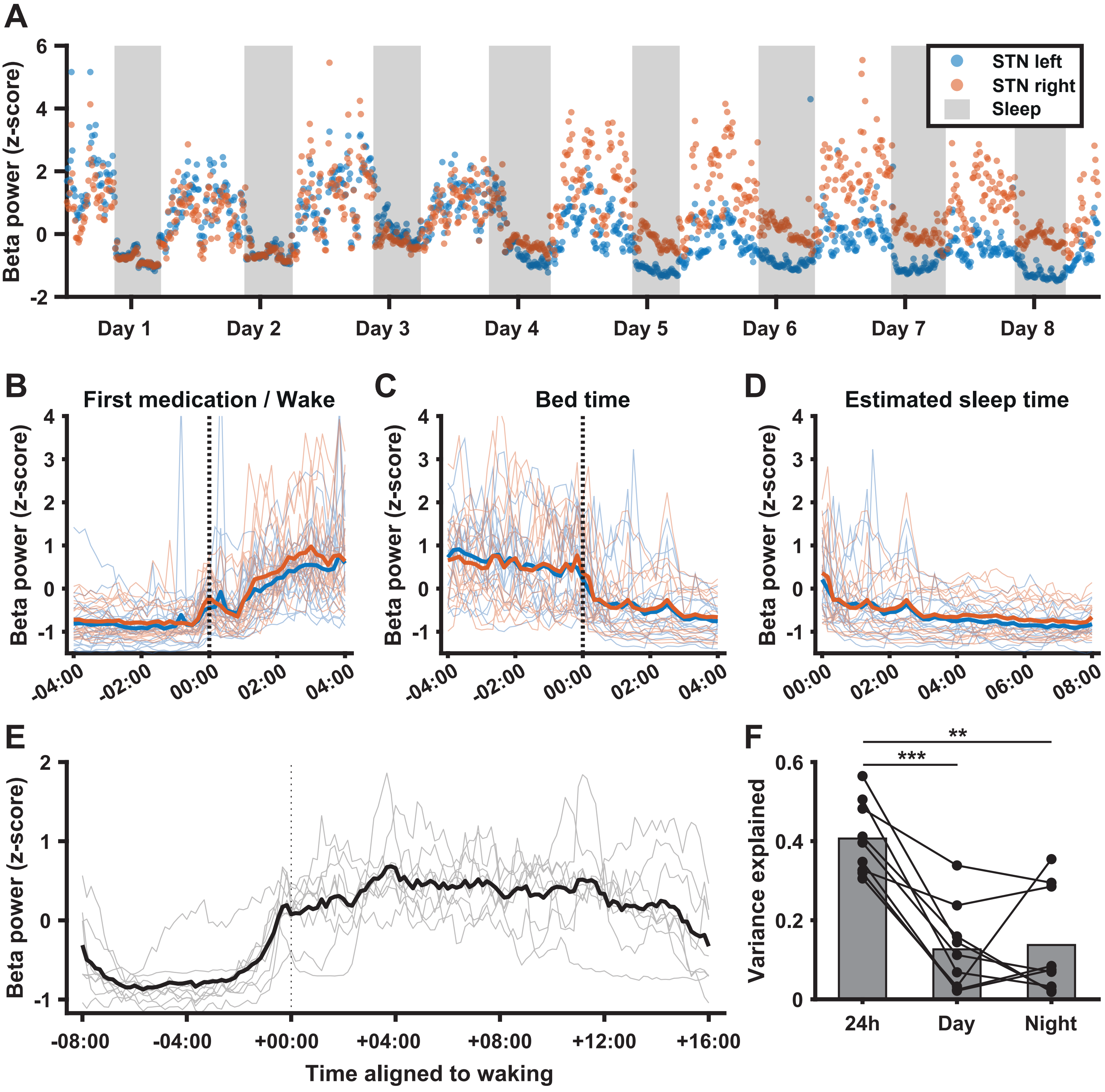

### Figure 3

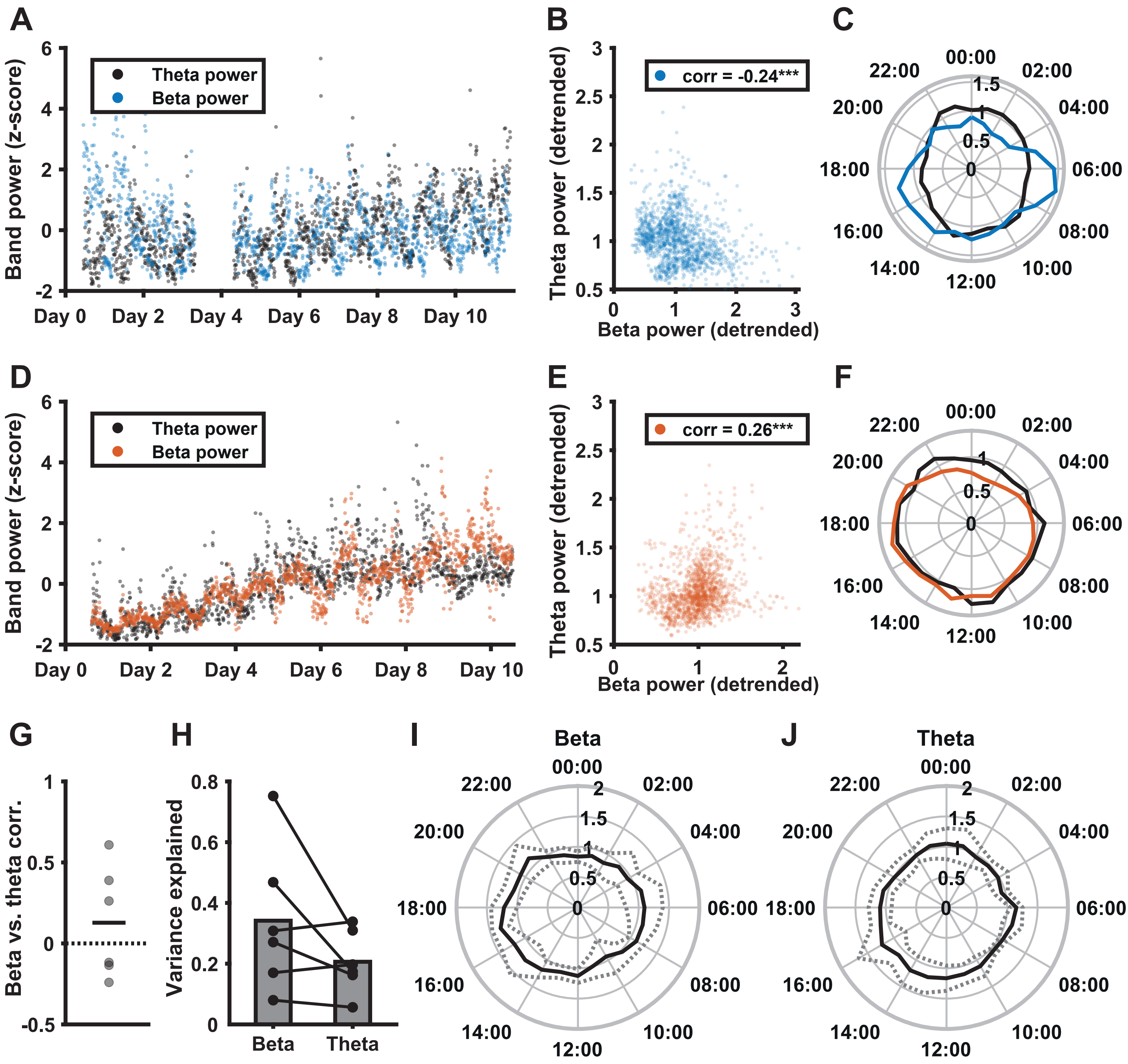

### Figure 4

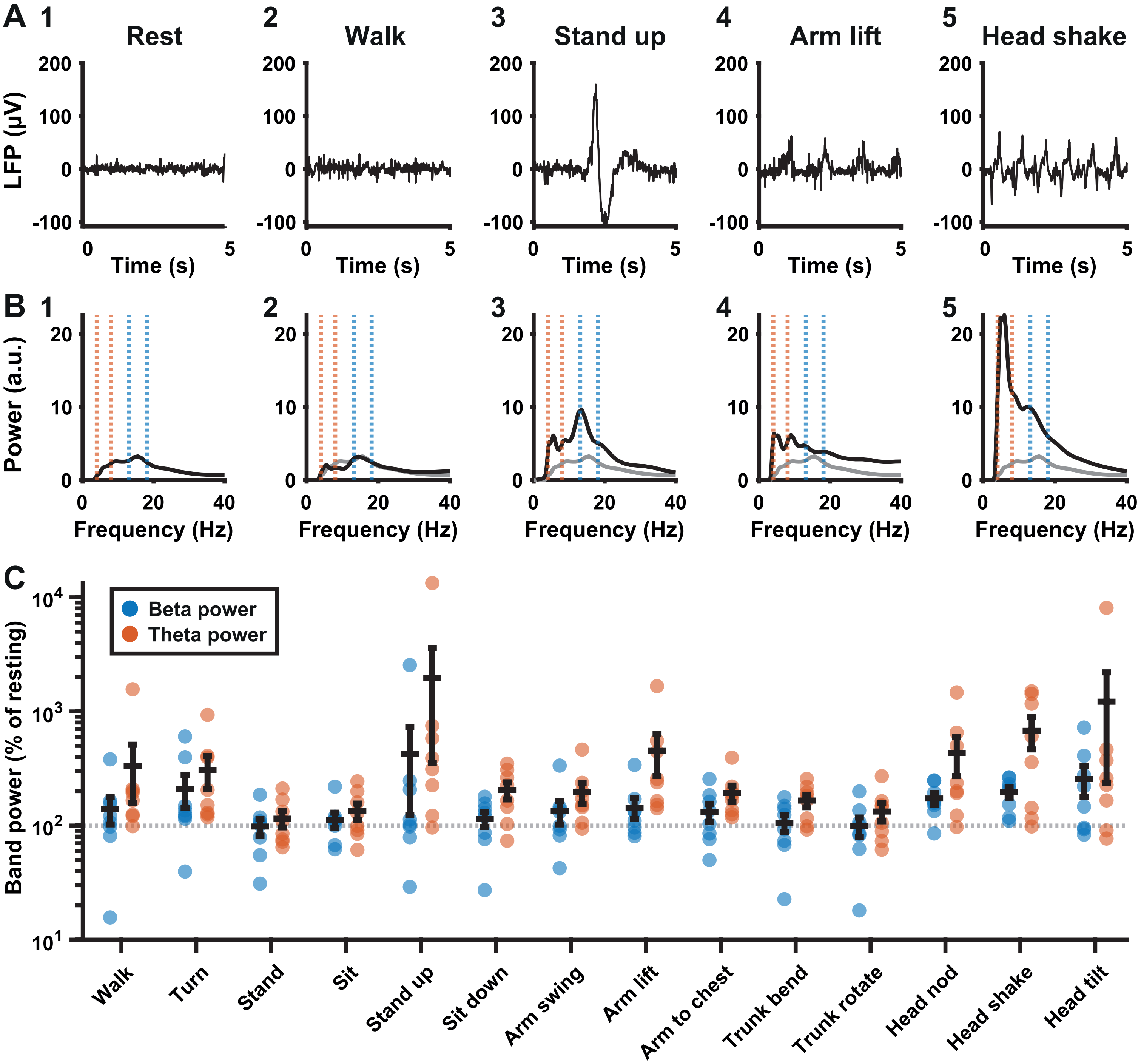

### Figure 5

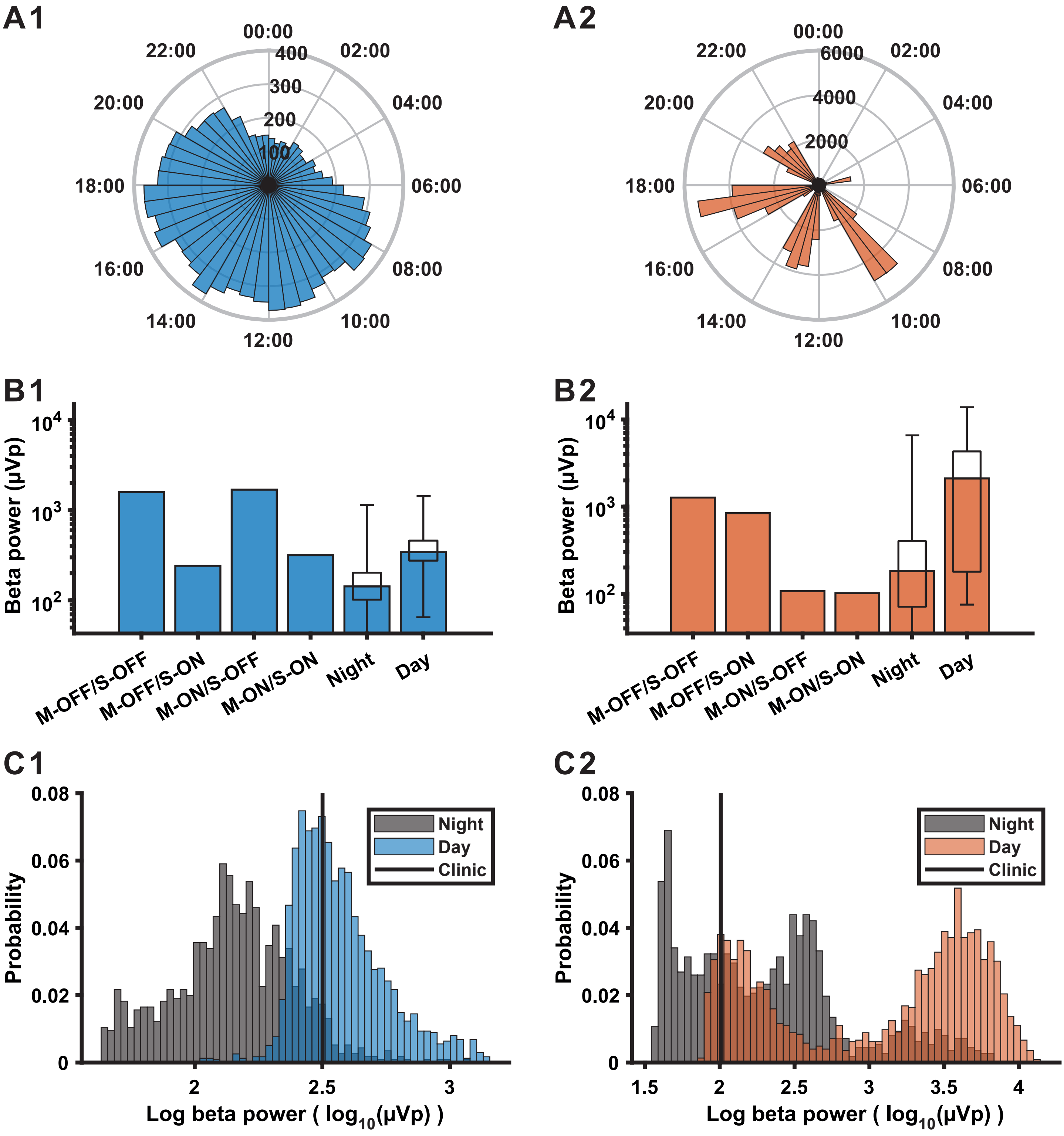

### Figure 6

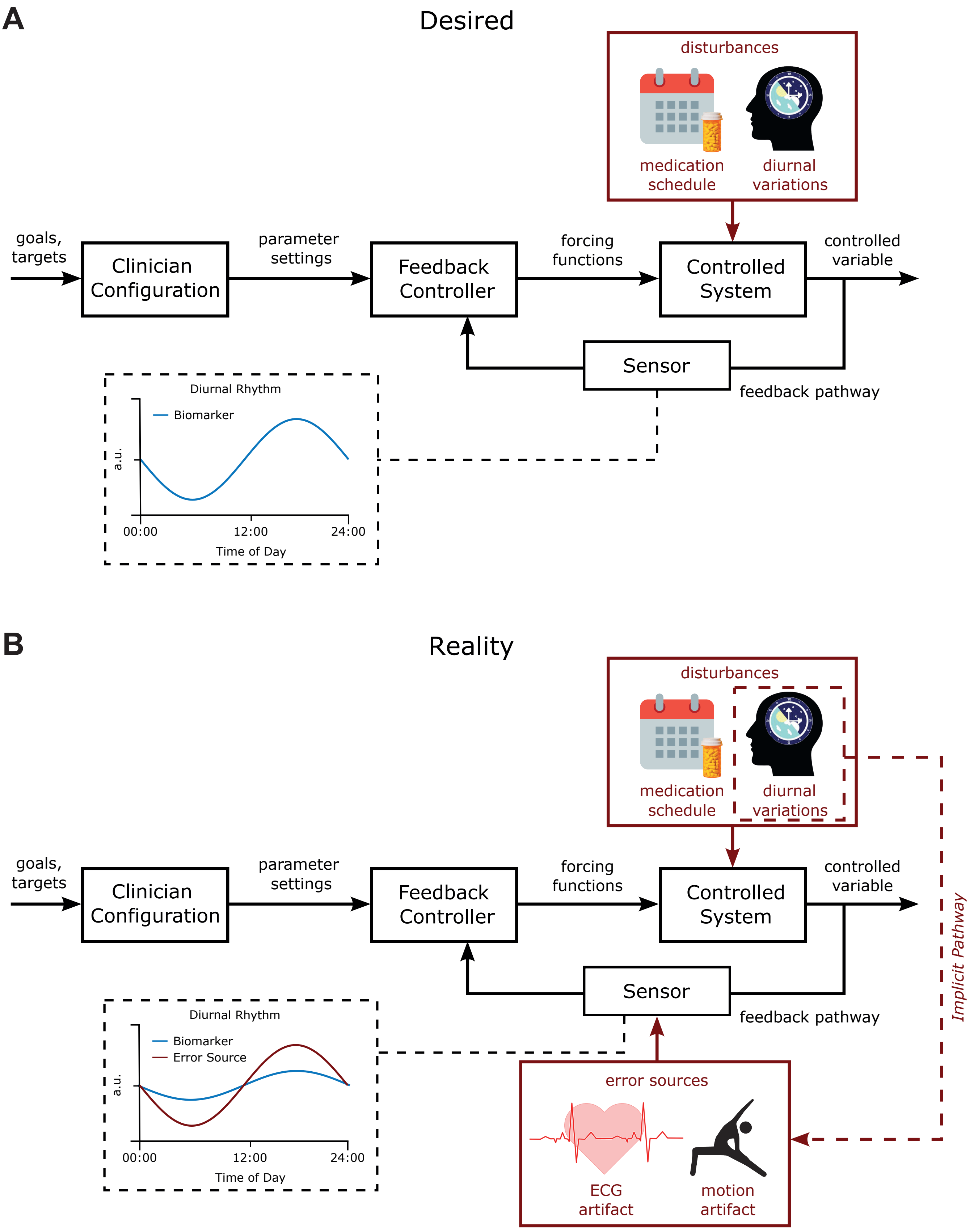

### Figure S1

#### Figure S1 - Patient #1 overview

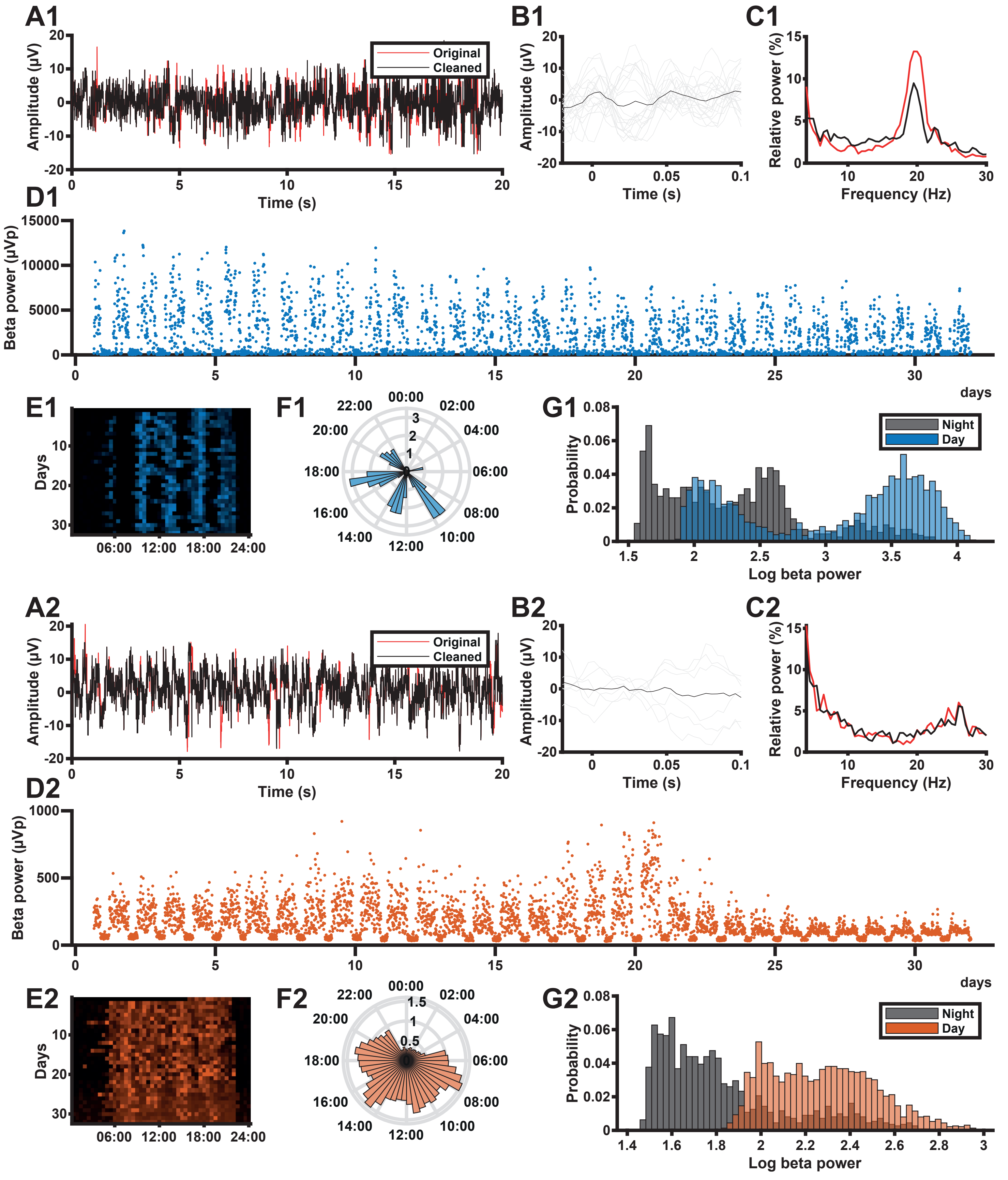

### Figure S2

#### Figure S2 - Patient #2 overview

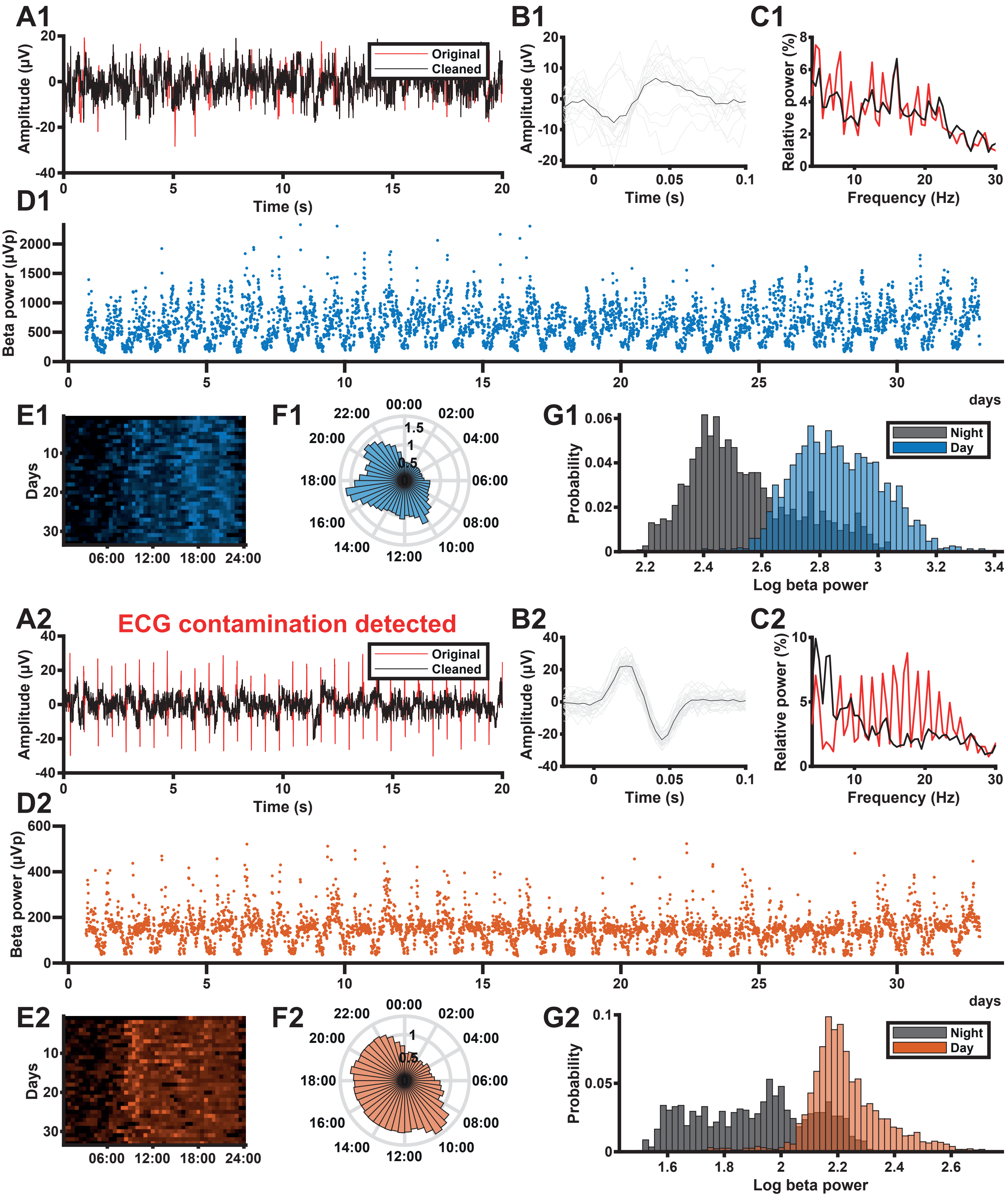

### Figure S3

#### Figure S3 - Patient #3 overview

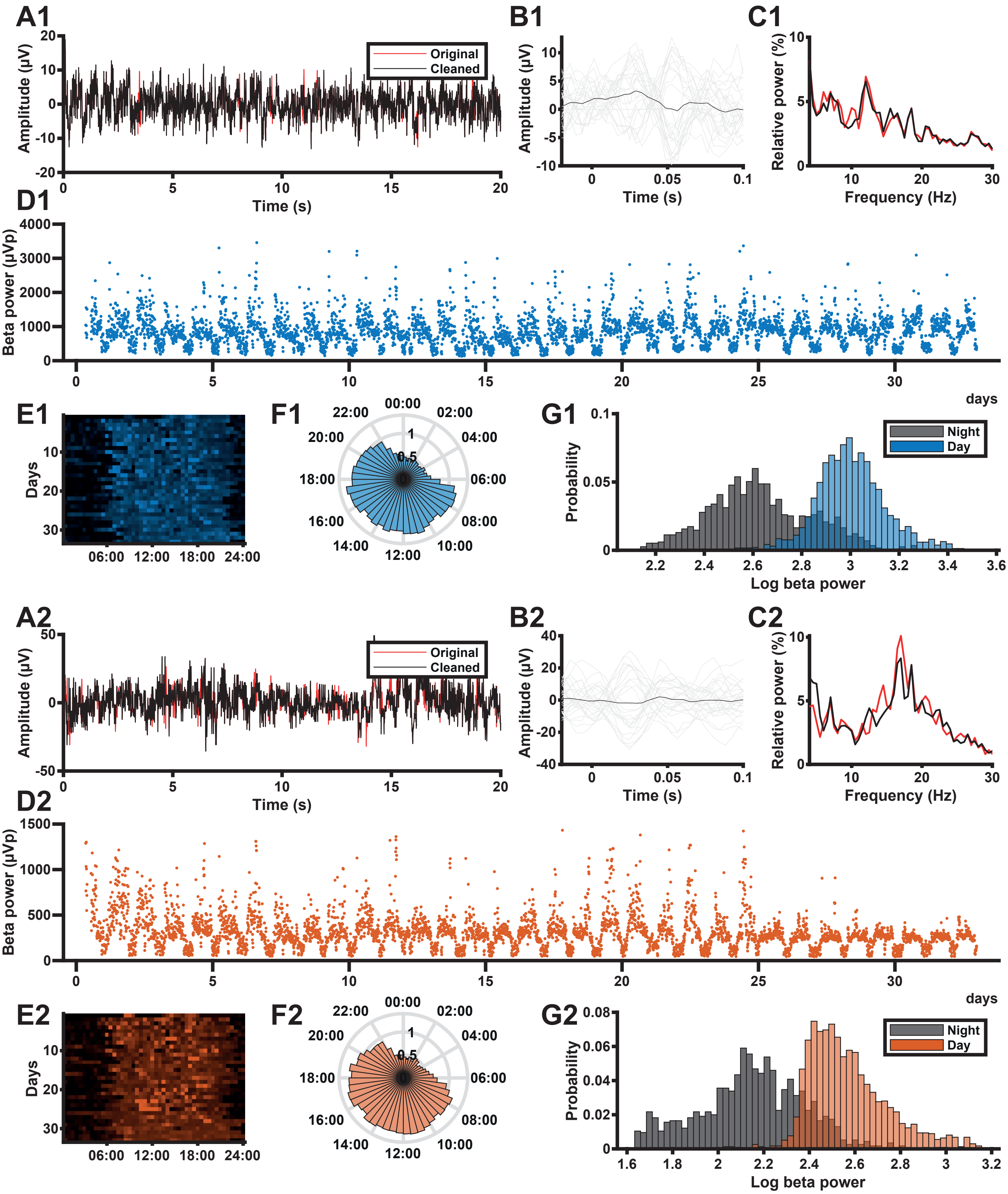

### Figure S4

#### Figure S4 - Patient #4 overview

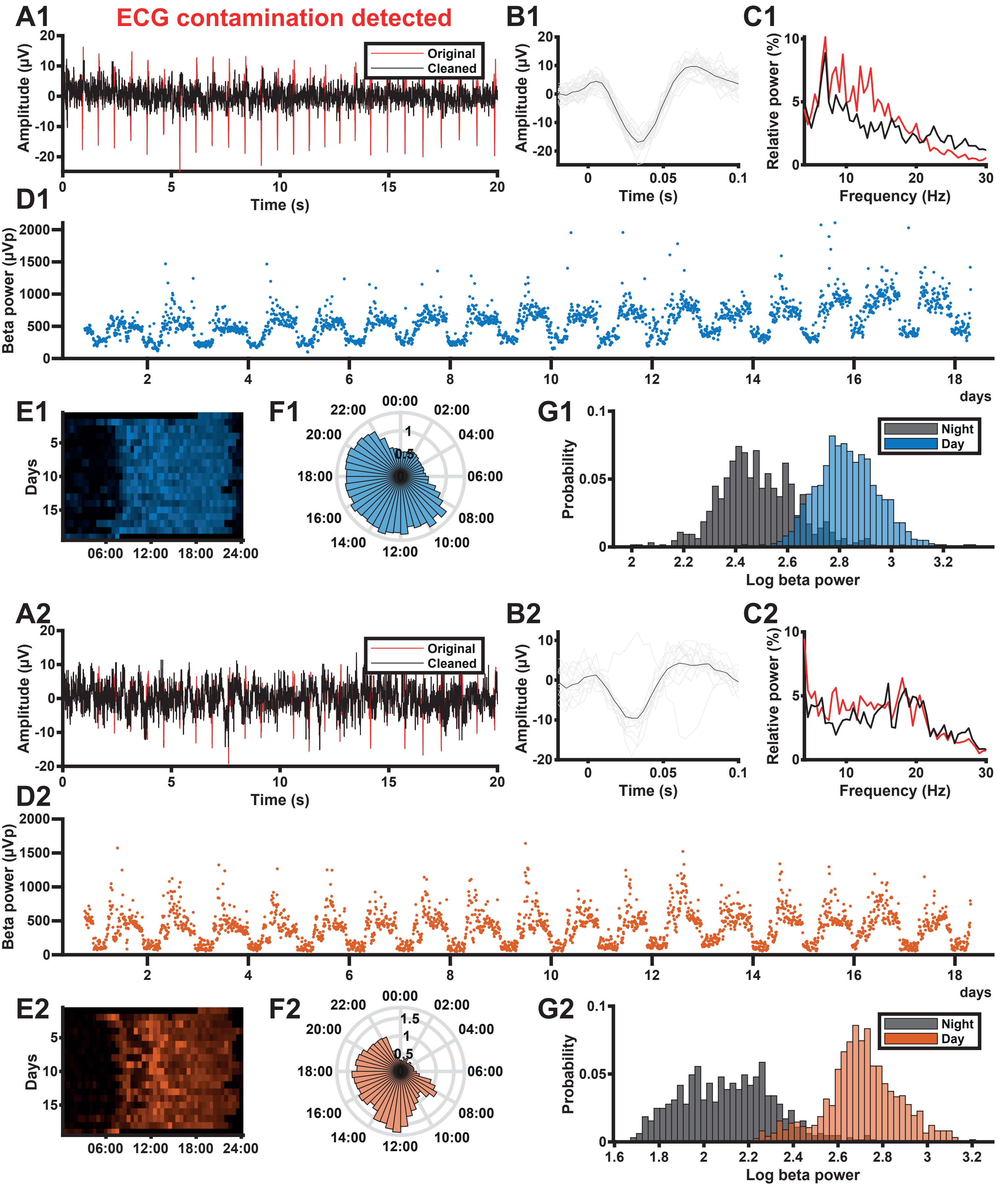

### Figure S5

#### Figure S5 - Patient #5 overview

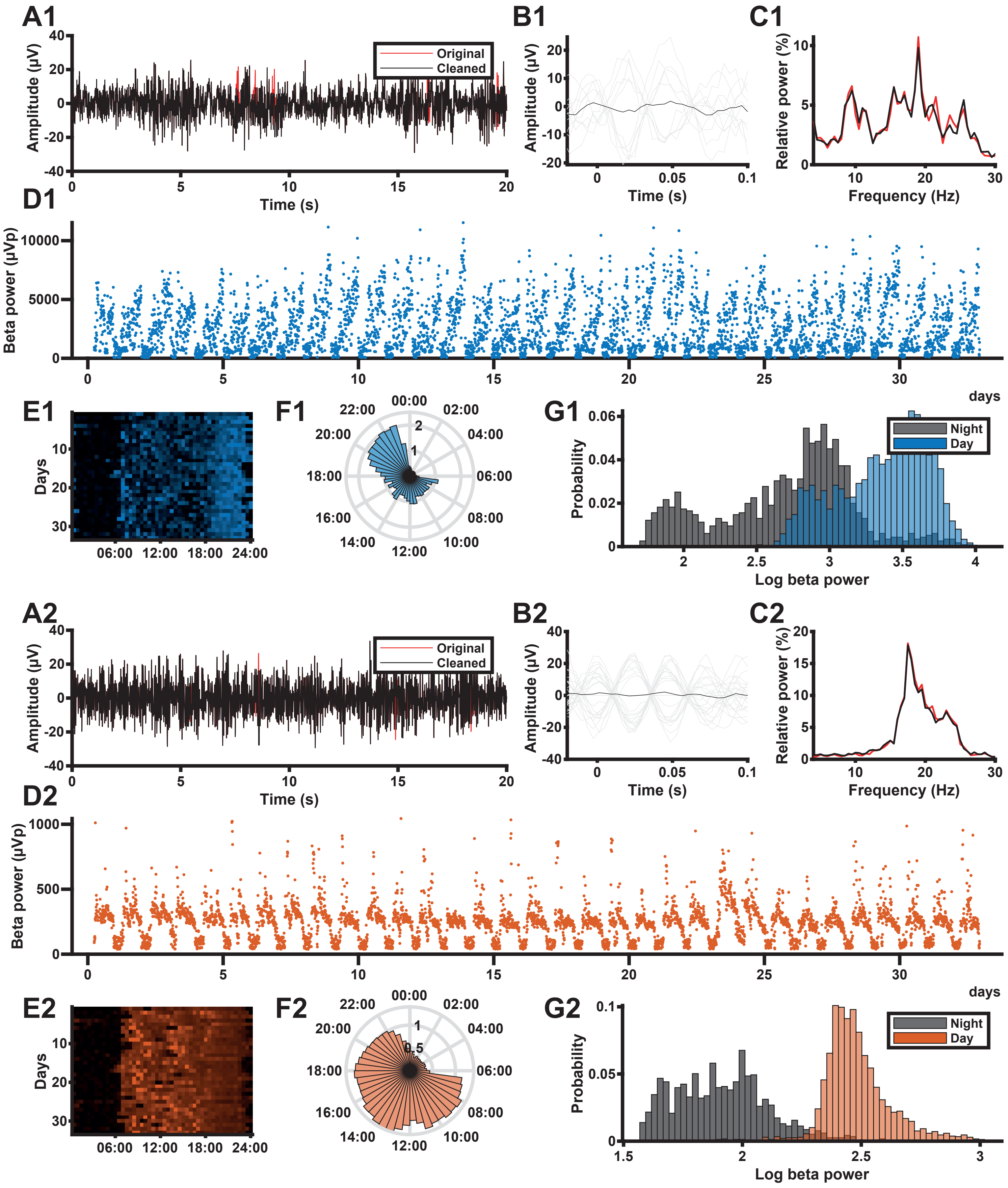

### Figure S6

#### Figure S6 - Patient #6 overview

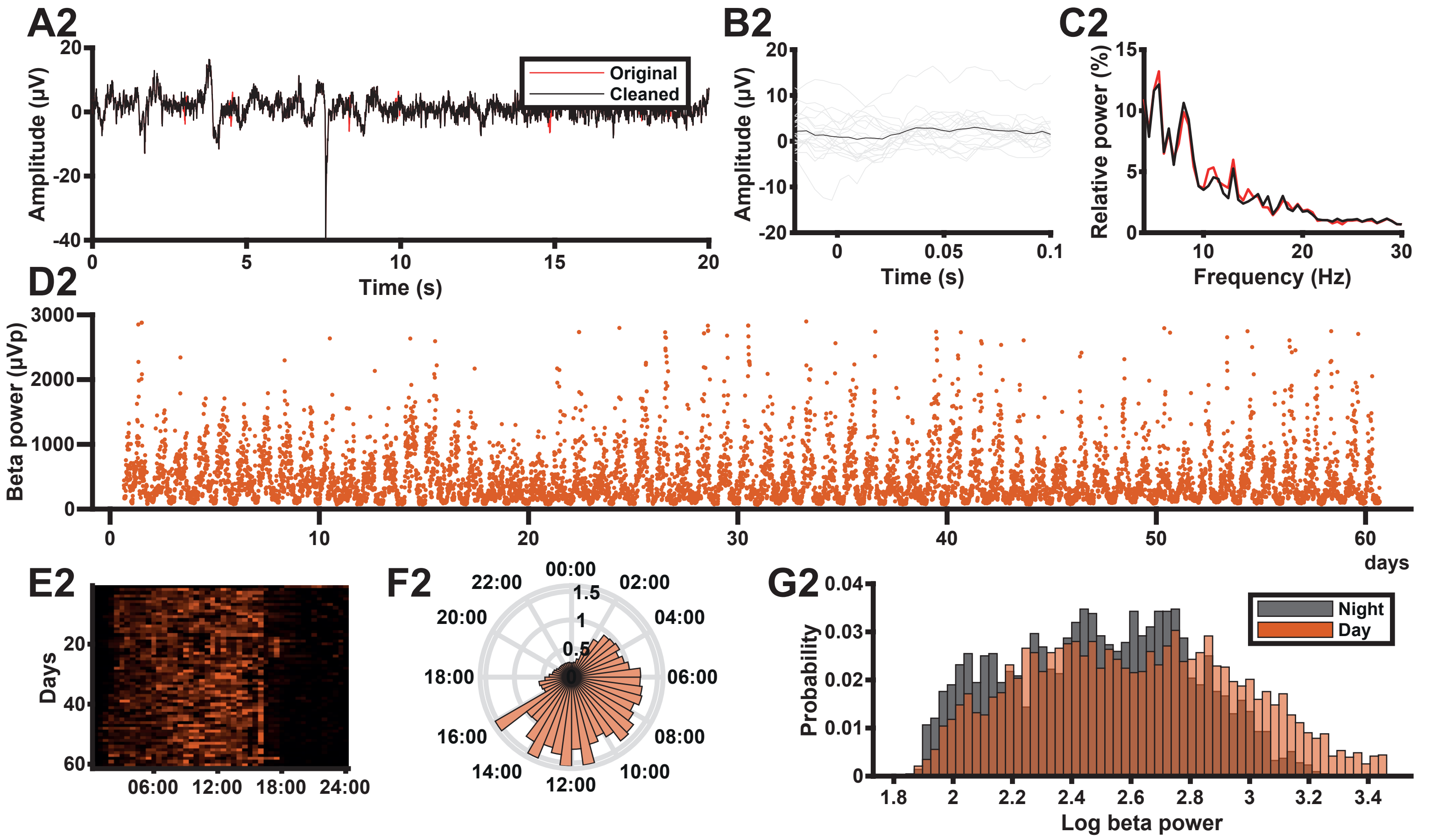
